# Targeting hepatitis B vaccine escape using immunogenetics in Bangladeshi infants

**DOI:** 10.1101/2023.06.26.23291885

**Authors:** Guillaume Butler-Laporte, Kathryn Auckland, Zannatun Noor, Mamun Kabir, Masud Alam, Tommy Carstensen, Genevieve L Wojcik, Amanda Y Chong, Cristina Pomilla, Janelle A Noble, Shana L. McDevitt, Gaby Smits, Susan Wareing, Fiona RM van der Klis, Katie Jeffery, Beth D Kirkpatrick, Sodiomon Sirima, Shabir Madhi, Alison Elliott, J Brent Richards, Adrian VS Hill, Priya Duggal, PROVIDE authors, Cryptosporidiosis Birth Cohort authors, Manjinder S Sandhu, Rashidul Haque, William A Petri, Alexander J Mentzer

**Affiliations:** Wellcome Centre for Human Genetics, University of Oxford, Oxford, United Kingdom; Lady Davis Institute, Jewish General Hospital, McGill University, Montréal, Québec, Canada; Division of Infectious Diseases, McGill University Health Centre, Montréal, Québec, Canada; International Centre for Diarrhoeal Disease Research, Bangladesh, Dhaka, Bangladesh; Wellcome Trust Sanger Institute, University of Cambridge, Hinxton, United Kingdom; Queen Mary University of London, London, United Kingdom; Department of Epidemiology, Johns Hopkins Bloomberg School of Public Health, Baltimore, Maryland, USA; Children’s Hospital Oakland Research Institute, Oakland, California, USA; Department of Pediatrics, University of California, San Francisco, California, USA; Media Labs, Inc. Alameda, CA, USA; National Institute for Public Health and the Environment, Bilthoven, The Netherlands; Microbiology Department, John Radcliffe Hospital, Oxford University NHS Foundation Trust, Oxford, UK; Department of Microbiology and Molecular Genetics, Vaccine Testing Center, University of Vermont College of Medicine, Vermont, USA; Groupe de Recherche Action en Santé (GRAS) 06 BP 10248 Ouagadougou, Burkina Faso; South African Medical Research Council Vaccines and Infectious Diseases Analytics Research Unit, University of the Witwatersrand, Johannesburg, South Africa; Department of Clinical Research, London School of Hygiene & Tropical Medicine, London, UK; Medical Research Council/Uganda Virus Research Institute and London School of Hygiene & Tropical Medicine Uganda Research Unit, Entebbe, Uganda; Department of Human Genetics, McGill University, Montréal, Québec, Canada; 5 Prime Sciences Inc, Montreal, Quebec, Canada; Department of Epidemiology, Biostatistics and Occupational Health, McGill University, Montréal, Québec, Canada; Department of Twin Research, King’s College London, London, United Kingdom; The Jenner Institute, University of Oxford, Oxford, United Kingdom; Department of Epidemiology and Biostatistics, School of Public Health, Imperial College London, United Kingdom; Department of Medicine, Infectious Diseases and International Health, University of Virginia School of Medicine, Charlottesville, Virginia, USA

**Keywords:** Genome-wide association studies, human leukocyte antigen, major histocompatibility complex, vaccination, hepatitis B virus, escape variants

## Abstract

Hepatitis B virus (HBV) vaccine escape mutants (VEM) are increasingly described, threatening progress in control of this virus worldwide. Here we studied the relationship between host genetic variation, vaccine immunogenicity and viral sequences implicating VEM emergence. In a cohort of 1,096 Bangladeshi children, we identified human leukocyte antigen (HLA) variants associated with response vaccine antigens. Using an HLA imputation panel with 9,448 south Asian individuals *DPB1*04:01* was associated with higher HBV antibody responses (p=4.5×10^−30^). The underlying mechanism is a result of higher affinity binding of HBV surface antigen epitopes to DPB1*04:01 dimers. This is likely a result of evolutionary pressure at the HBV surface antigen ‘a-determinant’ segment incurring VEM specific to HBV. Prioritizing pre-S isoform HBV vaccines may tackle the rise of HBV vaccine evasion.

**One-Sentence Summary:** Host genetics underlying hepatitis B vaccine response in Bangladeshi infants identifies mechanisms of viral vaccine escape, and how to prevent it.

## Main Text

Vaccination is the most effective prevention method against infections diseases and their sequelae. However, its success depends upon accurate targeting of the immune response against the pathogen and sustained immunogenicity. Hepatitis B virus (HBV) vaccines represent a prime example of this. Despite their success at reducing HBV-related disease burden worldwide, there is increasing evidence of vaccine escape mutations (VEMs) emerging because of variants in the “a”-determinant region of the surface antigen (HBsAg) gene. A recent study reported that 51% of GenBank Bangladeshi HBV isolates from 2005 to 2017 showed evidence of VEMs *1*. While selection bias may have inflated this estimate, robust evidence shows that vaccination campaigns lead to increasing VEMs prevalence *1–4*. In Taiwan, the prevalence of “a”-determinant VEMs increased from 7% to 28.1% following universal vaccination *5, 6*. In developing countries, where delays in vaccination program implementation may occur, a rise in HBV VEMs could hinder our best defense against infection. This is especially worrisome as HBsAg VEMs may lead to mutations in the reverse transcriptase gene, inducing antiviral treatment resistance *1*.

The primary HBV vaccine in worldwide use is the yeast recombinant HBV vaccine*7*, obtained from the *adw* subtype*8* of the HBsAg S isoform (fig. 1) *9–11*. This is now available in many combination vaccines used in national childhood vaccination strategies. A different HBV vaccine construct was developed as part of PreHevbrio (VBI Vaccines Inc., Cambridge, USA) which uses the full HbsAg protein (i.e. including the pre-S1 and pre-S2 segments) (fig. 1) and was approved for adult use in 2021. Pre-S1/pre-S2/S antigen vaccines have proven effective in inducing immunity even in non-responders to S isoform vaccines*12–17*. Non-response to HBV vaccines and its host factors are well described, and is largely explained by genetic variation in the human leukocyte antigen (HLA) gene cluster *18*. However, the specific components of this variation have not been defined across populations owing to both technical and population-specific challenges with this complex locus of the human genome.

**Fig. 1:**
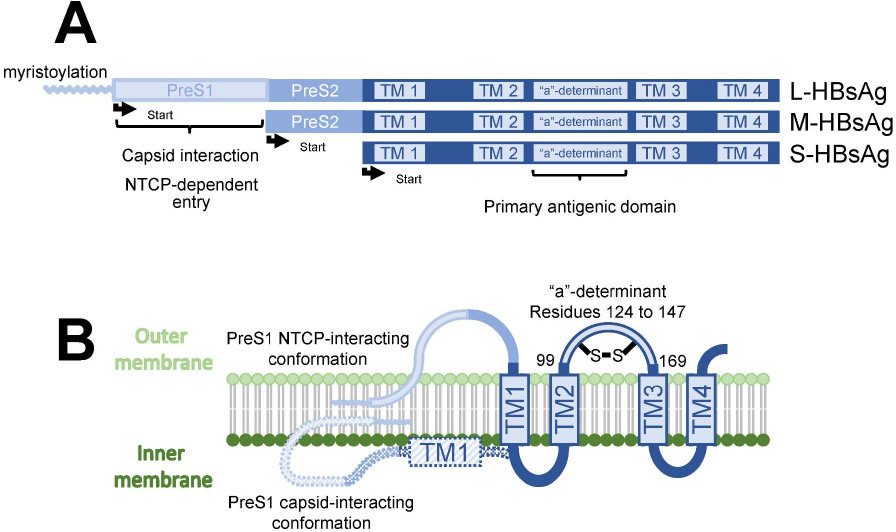
Schematic of the three HBsAg protein isoforms. **A)** The most restricted S-HBsAg isoform is used in yeast recombinant vaccine production. The pre-S1 segment is crucial for capsid interaction during virion assembly, and for hepatocyte viral entry in the first phase of HBV infection when it interacts with sodium taurocholate co-transporting polypeptide (NTCP). **B)** Representation of the two HBsAg conformations on the surface of HBV. The primary antigenic domain “a”determinant (where most known vaccine escape variants are found) is located between transmembrane domains TM1 and TM2, facing the outside of the virion. Figure adapted with permission from Vaillant, 2021(*63*).

The relationship between the emergence of HBV VEM and variable immunogenicity to HBV vaccines from HLA variants has not been well explored but given that class II HLA receptors are responsible for exogenous antigen presentation, there is likely a relationship between the two. Given that genetics can lead to actionable interventions when supported by replicable evidence*19–22*, it could guide vaccine development and prevent the rise of VEMs. Hence, there is an urgent need to improve our understanding of these mechanisms.

Here, we identified the genetic determinants of immune response to multiple vaccines including HBV in a cohort of Bangladeshi children for which antibody levels (a strong correlate of immunity) were measured at the same time following vaccination. We built the largest south Asian (SAS) genetic ancestry HLA imputation panel (at 2-field i.e. protein altering variants) using 9,448 whole-exome sequences from the UK Biobank (UKB) to fine-map HLA alleles and amino acid residues associated with antibody levels. We then used HLA receptor binding algorithms to show that antibody levels correlate with a participant’s HLA-DP receptors. Importantly, our results correlate well with epidemiological observation of HBV VEMs and with previously published randomized trials of HBV vaccination, suggesting a way forward to translate HLA association studies results into actionable therapeutic targets.

## Results

### Study cohort

The cohort consisted of a sample of Bangladeshi children from the PROVIDE (n=502) and the Cryptosporidiosis Birth Cohort (CBC) (n = 594) studies*23–26*. Both cohorts are extensively described elsewhere*23–30*. There were 534 females (n=236 for PROVIDE and n=298 for CBC) and 562 males (n=266 for PROVIDE and n=296 for CBC). Participants were genetically homogeneous (**fig. S1**) and were pooled for the rest of the analyses. Participants received vaccination against HBV, diphtheria, pertussis, tetanus, *Haemophilus influenzae* type B (HiB), measles, and rubella as per the national Bangladesh Expanded Program on Immunizations. Blood samples were collected from all the children at the age of 52 weeks for immunoglobulin G (IgG) measurements using validated multiplex immunoassays for the following antibodies: HBsAg, DT, tetanus toxin (TT), pertussis toxin (PT), pertactin (PRN, a *Bordetella pertussis* virulence factor), filamentous haemagglutinin (FHA, another *B. pertussis* virulence factor), HiB polysaccharide, and measles virus. Henceforth we refer to serological phenotypes using the acronyms listed above, even if referring to antibody levels (e.g. HBsAg instead of anti-HBsAg).

### Genome-wide association studies

After quality control (QC) and imputation with the TOPMed panel*31*, we used Regenie*32* to perform a genome-wide association study (GWAS) of each qq-normalized serological trait on 9,581,948 variants, adjusting for 10 genetic principal components, the cohort, and sex. The strongest association was found for HBsAg at chr6:33061456:C:CT (beta for C: 0.48, se: 0.04, p: 2.6×10^−28^, AF for C: 28.1%) (fig. 2). This variant was in a haplotype where the only two protein-coding genes are *HLA-DPA1* and *HLA-DPB1* (**fig. S2A**). These associations were observed in both CBC and PROVIDE (**fig. S3-4**).

**Fig. 2:**
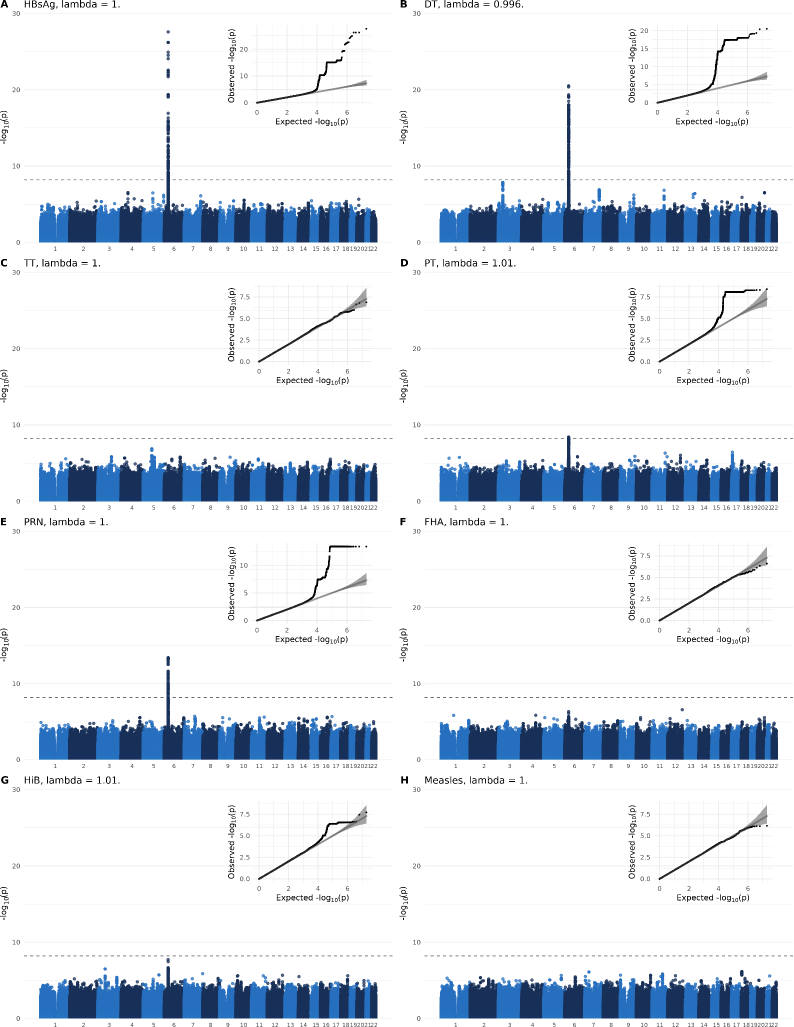
Manhattan and qq-plots of the single variants GWAS for each serology phenotype. The genome-wide significance line is set at 5×10^−8^/8. As shown by the genetic inflation factors (lambdas), there were no obvious signs of population stratification bias.

The second most significant association came at chr6:32466021:C:T for DT (beta for T: 0.35, se: 0.04, p: 3.0×10^−21^, AF for T: 46.7%). This variant lies in a complex HLA haplotype containing the *HLA-DRB1*, *HLA-DRB3*, *HLA-DRB4*, *HLA-DRB5*, *HLA-DQA1*, and *HLA-DQB1* genes (**fig. S2B**).

Lastly, the pertussis serologies also yielded genome-wide significant associations for two *B. pertussis* virulence factors: chr6:32602046:AAT:A for PT (beta for AAT: 0.24, se: 0.04, p: 3.8×10^−9^, AF for AAT: 32.7%), and chr6:32635710:T:C for PRN (beta for T: −0.29, se: 0.04, p: 4.0×10^−14^, AF for T: 48.3%). Both variants are also located in the *HLA-DR* and *HLA-DQ* haplotype above (**fig. S2C-D**).

### HLA imputation

Given our GWAS results, we proceeded with HLA allele association studies. To do this, we needed to impute HLA gene haplotypes for each participant. However, given that publicly available HLA imputation panels have a small number of Bangladeshi or SAS ancestry reference sequences, they have limited accuracy for our cohort.

Hence, we used the UKB’s whole-exome sequences (WES) and HLA allele calls*33* from 9,448 SAS ancestry participants to build a SAS HLA reference panel. Details are provided in the **Methods**. Briefly, after QC and merging of the UKB’s WES and whole-genome genotyping variants, we used a modified version of the HLA-TAPAS pipeline*34* to obtain a HLA reference panel at 2-field resolution, and impute HLA alleles for our cohort (see **data S1** for tag SNPs).

To check the accuracy of our results, we obtained independent HLA genotyping calls on 541 participants using a clinically validated 454 sequencing platform (Roche, Basel, Switzerland), using PCR-based single HLA exon calling methods. Concordance was over 90% in all genes except for *HLA-DRB3*, *HLA-DRB4*, *HLA-DRB5* (**table S1**), for which it was between 86.9% and 89.9%. The slightly lower concordance in those genes is expected given that they are not found in every individual, making it harder to distinguish a missing gene from a poorly imputed one. Importantly, we adjusted concordance values for the difference in technologies, providing *conservative* estimates of imputation accuracy (see **Methods**). Given the high concordance, we moved ahead with these for our HLA association studies.

### HLA allele association studies

We performed HLA allele association studies using the same serology phenotypes and analysis method as for the GWAS (**data S2**). This yielded five genome-wide significant HLA allele associations (**table S2**). The most significant observed association was *DPB1*04:01* with HBsAg (beta: 0.491, se: 0.04, 95% CI: 0.40 – 0.56, p: 4.5×10^−30^). The other four were observed with DT : *DQA1*02:01* (beta: 0.29, se: 0.04, 95% CI: 0.21 – 0.37, p: 1.9×10^−12^), *DQB1*06:01* (beta: −0.31, se: 0.05, 95% CI: −0.40 – −0.22, p: 2.5×10^−11^), DRB1*07:01 (beta: 0.29, se: 0.04, 95% CI: 0.21 – 0.38, p: 1.2×10^−12^), and *DRB4*01:03* (beta: 0.26, se: 0.04, 95% CI: 0.17 – 0.34, p: 1.2×10^−9^).

We also performed conditional analyses on alleles associated with DT to determine whether they were independent (**table S2**, **data S3**). *DRB1*07:01* and *DQA1*02:01* alleles were not independent, as expected by their high linkage disequilibrium (LD) (r^2^ = 0.99, **data S4**). The signal from *DRB4*01:03* was also lessened when conditioned on either of these two alleles, also expected from their mild LD (r^2^ = 0.33). However, the association between DT and *DQB1*06:01* was preserved when conditioning on the other alleles. Given the complex LD between HLA alleles at this locus, we also used HLA dimers-based tests to better dissect the HLA-DQ alleles signal (see **Supplementary Text** and **data S5**).

### Amino acid residue association studies

To understand the components of each HLA receptor dimer associated with antibody levels, we mapped each participant’s HLA allele to their amino acid residues and performed amino acid association analyses (**data S6**). Results matched those found in the HLA allele associations above. That is, for HBsAg and DT, all HLA genes found in allele association tests above also carried amino acid residues that were significantly associated with the phenotypes (fig. 3). For example, residues 85G, 86P, and 87M of *HLA-DPB1* were the most associated with HBsAg (beta: 0.33, se: 0.04, p: 4.89×10^−17^, 95% CI: 0.25 to 0.40), and residues 98K and 104S were the most associated with DT (beta: −0.32, se: 0.04, p: 2.86×10^−18^, 95% CI: −0.40 to −0.25). Importantly, most amino acid uncovered were in the HLA receptors’ peptide binding groove (fig. 3 and **fig. S5**), in keeping with the vaccine’s expected biological mechanism.

**Fig. 3:**
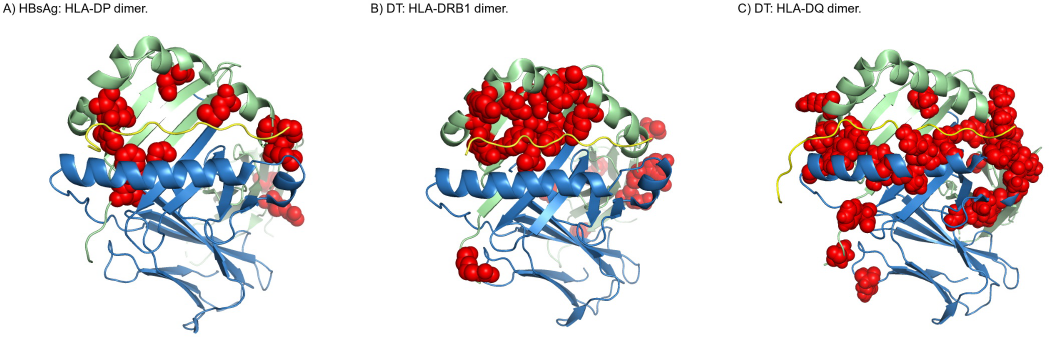
Visualization of amino acid residues (in red) associated with each serology phenotype. For each dimer, the blue chains represent the alpha subunits (i.e. DPA1, DRA, and DQA1, from left to right), the green chains represent the beta subunits (i.e. DPB1, DRB1, and DQB1, from left to right), and the yellow chains show the location of a given epitope in the dimers’ peptide binding grooves. See **fig. S5** for similar visualization for DT and HLA-DRB5, and for PRN and HLA-DQ.

### HBV and DT in-silico HLA dimer binding to disentangle HLA linkage disequilibrium

We used NetMHCIIpan*35* to independently validate our HLA associations. We assessed the predicted binding affinities of each HLA haplotype against epitopes from 50 HBsAg and 12 DT protein sequences on Uniprot*36*. This was done for *HLA-DPA1-DPB1* haplotypes for HBsAg, and for *HLA-DRB1* and *HLA-DQA1-DQB1* for DT. NetMHCIIpan is a validated neural network that predicts binding affinity for an HLA dimer and a 15-mer epitope of any antigen. It is therefore not affected by linkage disequilibrium.

For HBsAg, the *DPB1*04:01* containing HLA-DP receptor dimers were predicted to bind the S protein isoform of the HBsAg protein with greatest affinity compared to dimers containing other HLA-DPB proteins in our cohort (fig. 4). The S isoform of the HBsAg is contained in the yeast recombinant vaccines given to our participants (fig. 1). Hence, both genetic association studies and *in-silico* binding affinity results agreed.

**Fig. 4:**
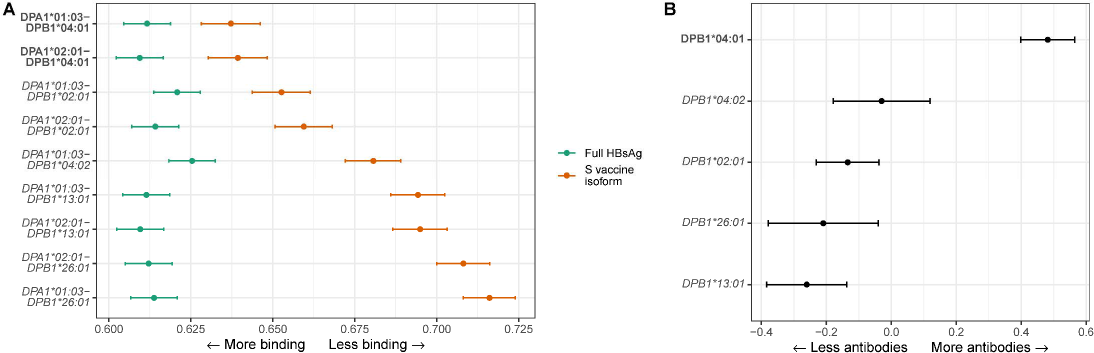
**A)** *in-silico* binding results from NetMHCIIPan. Points are the effect coefficients from beta regression run on the rank outputs of predicted binding of all 15-mers derived from 50 HBsAg protein sequences. In green, results are shown for the full HBsAg protein sequence. In orange, results are shown from an analysis restricted only to 15-mers within the S isoform of HBsAg. The S isoform is the one used in making yeast recombinant vaccines, such as those given to our participants. Coefficients and 95 % CIs are shown. The more they are positioned to the left, the higher average *in-silico* binding across the antigen. Dimers in bold contain the *DPB1*04:01* allele, to better compare with the HBsAg antibody association tests in the right panel. **B)** Results from the HLA allele association studies using linear regression with antibody responses and imputed HLA alleles from the Bangladeshi datasets. Again, coefficients and 95% CIs are shown. Values to the right on the x-axis indicate greater HBsAg antibody measures. The *DPB1*04:01* allele is shown in bold to better compare with the *in-silico* results in the left panel

For DT, *in-silico* binding also matched the genetic association tests (spearman correlation rho = - 0.50, p = 0.03), and the HLA-DR allele associated with the highest level of DT (DRB1*07:01) was also the best DT proteins *in-silico* (**fig. S6**). However, we see instances where LD may have biased our genetic association tests. For example, the DQA1*01:03−DQB1*06:01 dimer was associated with the second lowest antibody levels but was the third best *in-silico* DT proteins binder. *DQA1*01:03* is in moderate LD with *DRB1*15:02* (r^2^ = 0.39), while *DQB1*06:01* is in moderate LD with both *DRB1*15:01* (r^2^ = 0.25) and *DRB1*15:02* (r^2^ = 0.27). These all showed low antibody levels and poor *in-silico* binding. Hence, binding affinity disentangled the LD patterns at the HLA locus.

### Pre-S isoforms, a-determinant VEMs, and HBV vaccine design

Given the correlation between the HLA association results and the *in-silico* binding results, we investigated the reasons for these differences between *HLA-DPB1* alleles. To do this, we plotted the difference in predicted binding affinity between the best performing *HLA-DPB1* allele (*DPB1*04:01*) and the worst performing allele (*DPB1*26:01*), using *DPA1*01:03* as their common pairing subunit (fig. 5).

**Fig. 5:**
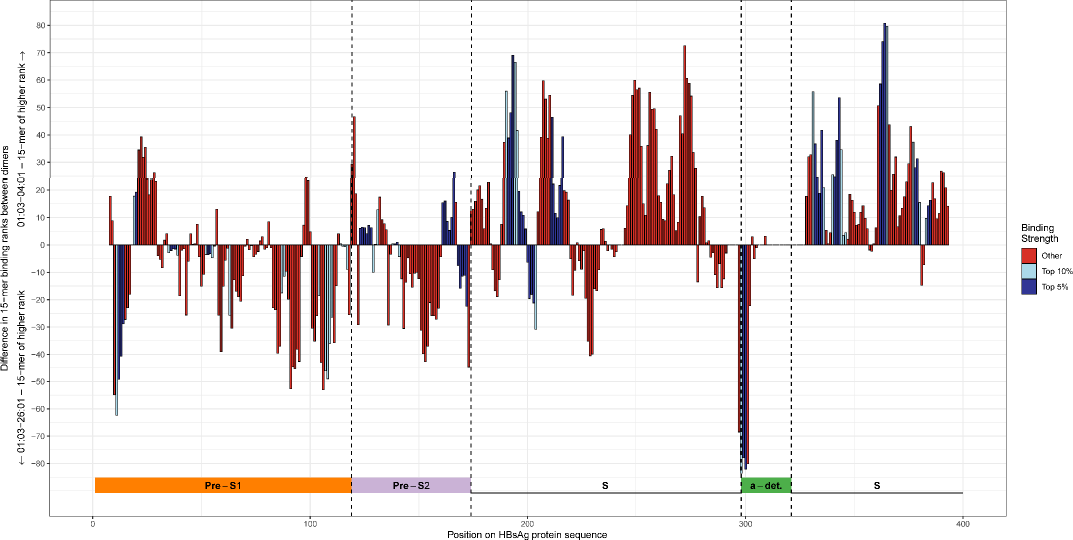
Difference in HBsAg binding affinity between the dimer with highest predicted binding affinity to HBsAg peptides (DPA1*01:03-DPB1*04:01) and the dimer with the lowest (DPA1*01:03-DPB1*26:01). The x-axis represents bins of 1 amino-acid across the entire HBsAg sequence. The y-axis represents the difference in binding affinity between the top-binding 15-mers binding in this region for both dimers. Positive values indicate greater binding for DPA1*01:03-DPB1*04:01 than for DPA1*01:03-DPB1*26:01. Negative values indicate the opposite. The colours of the vertical bars refer to the rank of affinity of binding of the 15-mer to either DPA1*01:03-DPB1*04:01 (positive values) or DPA1*01:03-DPB1*26:01 (negative values). Small blue bars mean that both dimers are predicted to bind with high affinity (top 10% quantile) at this position. Long red bars mean that the difference in dimer binding to 15-mer is predicted to be large, but weak for both. A long dark blue bar means that the expected binding difference is large and that one of the dimers bind strongly to the 15-mer. For example, 15-mers around position 300 of the HBsAg protein are expected to bind strongly to DPA1*01:03-DPB1*26:01 and to bind much better than to DPA1*01:03-DPB1*04:01. Bottom colored horizontal bar indicates the different section of the HBsAg protein.

There were two striking observations. First, DPB1*04:01 bound to more 15-mer epitopes from the HBsAg S isoform than DPB1*26:01. However, the pre-S containing isoforms contained a similar number of strongly binding epitopes for both DPB1 proteins. To confirm this observation, we fitted a beta regression to the *in-silico* results for the full pre-S containing HBsAg protein (fig. 4A). This showed a large difference in binding affinity between S and pre-S containing HBsAg isoforms, and an attenuated difference between HLA-DP dimers.

Second, the only region of the HBsAg S isoform that binds to DPB1*26:01 better than DPB1*04:01 was between amino acids 124 to 127. These residues are at the start of the well-known “a”-determinant of HBsAg. The a-determinant is of great clinical importance, as variants in this region lead to HBV strains which can overcome vaccines. In particular, mutations R122K, I126T, and P127L are associated with HBV VEMs, especially in Bangladesh*1–4*, and worsened the *in-silico* binding strength at DPB1*26:01 (**data S7**).

Hence, difference in epitope binding at the a-determinant between the different HLA-DP dimers is associated with VEMs. These VEMs worsen binding with a HLA protein (DPB1*26:01) already lacking other binding target in the S HBsAg isoform and showing decreased antibody response following immunization. More specifically, a single amino acid polymorphism at the a-determinant is the only barrier to VEMs in carriers of non-*DPB1*04:01* alleles, providing a potential HBV VEMs selection mechanism. Moreover, and concordant with the randomized trials showing that pre-S isoform vaccines can “salvage” non-responding patients, our data provides support the use of such vaccines in populations where HBV mutations are prevalent.

### HBsAg antibody level findings replication in the VaccGene cohort of African genetic ancestry participants

We replicated our results in an independent cohort of 2,499 infants from the VaccGene cohort in three African countries: Burkina Faso, South Africa, and Uganda*37*. All participants were vaccinated and genotyped using a similar methodology as ours. *DPB1*04:01* was also strongly associated with higher antibodies to HBsAg (beta = 0.27, se = 0.08, p = 6.3 x 10^−4^), but was much less frequent than in our cohort (AF = 4.3% in VaccGene vs 31.2% in the Bangladeshi cohort) (**table S3**). In VaccGene, the most significantly associated *HLA-DPB1* allele was *DPB1*01:01*, associated with a worse immunization outcome (beta = −0.23, se = 0.04, p = 1.1×10^−10^). A similar result was found in our Bangladeshi cohort (beta = −0.44, se = 0.13, p = 6.9×10^−4^), again with a large difference in allele frequencies (AF = 30% in VaccGene vs 2.2% in the Bangladeshi cohort). Hence, despite large differences in allele frequencies, our *HLA-DPB1* results are independently validated.

We performed the same binding affinity analysis as above to see if the pre-S/S isoform results observed with *HLA-DPB1* still held with *HLA-DQA1-DQB1* and *HLA-DRB1*. Again, we found that the binding affinity results mirrored the genetic association studies (**fig. S7-10**), and most importantly, that 15-mers from the pre-S segment of HBsAg were predicted to improve binding affinity for all alleles of *HLA-DQB1* and *HLA-DRB1*.

## Discussion

In this study, we built the largest SAS ancestry HLA imputation panel ever reported to find strong evidence that children from Bangladesh carrying specific HLA alleles mount a greater antibody response to certain vaccines. Specifically, we found that *DPB1*04:01* was associated with a stronger response to HBV vaccination, and that *DRB1*07:01* and *DQA1*01:03-DQB1*06:01* were associated with a stronger response to diphtheria vaccination. We also found evidence of a biological mechanism involving the HLA epitope peptide binding grooves, identified through the genetic association analysis, and supported by further *in-silico* analysis. Lastly, and more importantly, we showed that for the routinely administered S isoform HBV vaccines, the HLA alleles associated with worse post-vaccination antibody responses differed from *DPB1*04:01* by their stronger binding to HBsAg a-determinant epitopes, and that this binding was decreased by well-known VEMs. Since DPB1*04:01 can bind strongly to other epitopes from the S isoform of HBsAg, salvaging its binding to HBsAg even in the presence of a-determinant VEMs, we hypothesize that the combination of poorer antibody response and low VEMs threshold for the other DPB1 alleles could help explain the appearance of mutants at the a-determinant. These results highlight the potential use of pre-S HBsAg isoform HBV vaccines as a therapeutic option to prioritize in VEMs endemic areas, since non-DPB1*04:01 dimers are expected to bind with higher affinity to pre-S epitopes. Independent replication in VaccGene also supported the use of pre-S HBsAg isoform vaccines. In support of our results and conclusion, pre-S vaccination has already been demonstrated to induce effective immune responses in individuals classified as non-responders to yeast recombinant S isoform vaccine in multiple randomized trials*12–17*.

More generally, this study shows how to use orthogonal evidence to untangle the complex web of HLA allele associations in human diseases and conditions which limits the translation of HLA genetic results to therapy. Multiple factors limit interpretation of HLA association studies including reliance on European genetic ancestry over-represented imputation panels, high LD at the HLA locus, high pleiotropy of HLA alleles, and the difficulty in bridging HLA allele association results to their biological mechanism. These make any therapeutic target inference difficult. In this study, we tackled these problems by using an imputation panel that matched our study population, and by triangulating independent genetic, *in-silico*, and epidemiological data. Given the explicit knowledge of antigens driving HLA associations with infectious antigenic stimulation, this approach may lead to greater understanding of other HLA associated traits. For vaccination and therapies acting on the HLA epitope presenting pathway, therapy development could be influenced by knowing the HLA dimer binding affinity and its allele frequency in the target population.

Nevertheless, this study has limitations. First, while we found a well-reported “hotspot” of HBV mutation at amino acid 126 of the HBsAg S isoform, we did not find evidence that the G145R HBV VEMs would be preferentially selected. The G145R polymorphism is another well reported VEM *38*. However, it is believed G145R modifies the HBsAg three-dimensional conformation*39*, rather directly decreases its binding affinity to HLA dimers or antibodies. Our methods did not assess differences in protein conformation. Nevertheless, our results concerning the use of pre-S containing HBV vaccines has been confirmed through multiple trials in non-SAS cohorts with likely different genetic architectures, and our main conclusion still holds. Second, we used *in-silico* results to study binding strength of different antigens and their epitopes. While NetMHCIIpan is a validated well-performing algorithms that has shown great reliability in identifying strong HLA dimer binders, it is not the gold standard method to determine binding affinity. However, the fact that its results mirrored genetic associations and highlighted areas of high mutation risk independent of epidemiological data reinforces our findings. Our negative control results for DT (see **Supplementary Text**), where the *in-silico* locus of VEMs led to decreased pathogenicity, are also reassuring. Ultimately, confirmation of this work will require increased HBV case surveillance and linkage to sequencing and vaccination history, but these are challenging especially in low-to-middle income settings where VEM emergence is more likely.

In conclusion, in this Bangladeshi children cohort, orthogonal sources of genetic, *in-silico*, and epidemiological data showed that the *DPB1*04:01* allele improved response to the regular yeast recombinant S isoform HBV vaccine. We find genetic pressure on HBV to induce VEMs at the a-determinant due to human HLA alleles. Trials on pre-S containing HBV vaccines should be prioritized to limit the future rise of HBV VEMs in endemic regions. Methods used here are relevant for other vaccines where human genetic diversity underlies immune response.

## Methods

### Study cohorts

The participants were recruited from two separate vaccination cohorts*23, 24*: 1) the Performance of Rotavirus and Oral Polio Vaccines in Developing Countries (PROVIDE) study*25*, and 2) the Cryptosporidiosis Birth Cohort*26* (CBC). The CBC cohort was designed to study the effect of different gastrointestinal pathogens on child growth, while the PROVIDE cohort was an interventional 2×2 factorial design trial or oral rotavirus and polio vaccine (ClinicalTrials.gov registration no. NCT02764918). However, all participants were vaccinated per the national Bangladesh Expanded Program on Immunizations. This included the following: 1) pentavalent vaccine (diphtheria/pertussis/tetanus, *adw* hepatitis B, and *Haemophilus influenzae* type B) at 6, 10, and 14 weeks of age, 2) bivalent Measles-Rubella vaccine at 40 weeks of age, and 3) the monovalent measles vaccine at 65 weeks of age.

### Serology

Plasma samples were obtained at an approximate 52-week age point. The assays were described in detail elsewhere*37*. Briefly, three validated multiplex immunoassays were used to measure antibody concentrations against several vaccine antigens. This method measures total IgG against each respective antigen including functional (e.g. neutralizing) as well as non-functional antibodies. Antibodies against DT, TT, pertussis toxin (PT), pertactin (PRN), filamentous haemagglutinin (FHA), and MV were determined in the MDTaP assay, which is a combination of two previously described assays*40, 41*. Antibodies against Hib polysaccharide were determined in the HiB assay*42*. For MV and DT the correlation of the multiplex immunoassay to gold standard functional assays is high*43, 44*. The final concentration of bound antibody was calculated by determining the median fluorescence intensity of the antigen-specific beads and using diluted standards to calculate the concentration in international units for each antigen. Hepatitis B surface antigen (HBsAg) responses were measured using the anti-HBs kit on the ABBOTT Architect i2000 using recommended protocols (Abbott Laboratories, Chicago IL, USA).

### Genotyping

All participants were genotyped using the Multi-Ethnic Genotyping Array (Illumina Inc., San Diego, USA). The quality control steps (QC) were as follows: we removed all samples with a call rate < 97% with heterozygosity more than 3 standard deviations away from the mean, whose genetic sex did not match their reported one (threshold F > 0.8 for males and F < 0.2 for females), or with identity-by-descent > 0.9. We excluded genetic variants with a call rate < 97% or with Hardy-Weinberg equilibrium (HWE) p-value > 5×10^−8^. For males, we did not remove chromosome X variants based on HWE. After QC, we imputed additional genetic variants with the TOPMed imputation server*31* (August 8, 2022). Finally, we removed all variants with imputation r^2^ < 0.6 and minor allele frequency (MAF) < 0.5%.

### HLA imputation panel

We used a combination of the UKB’s hard-called genotype array data*45*, WES variants, and HLA allele calls from WES*46* of 9448 unrelated south Asian genetic ancestry participants in the UKB*33* in order to build a HLA reference panel at 2-field resolution (details of the HLA calling algorithm and QC are described elsewhere in full*33*).

First, we combined the UKB’s genotype array hard called variants with the WES calls to form the genetic variant backbone of our reference panel. We only used genotyped hard calls with HWE p-value > 5×10^−8^, and only used WES calls with HWE p-value > 5×10^−8^, average genotype quality of 10 or more, average depth of 10 or more, a minor allele count of 2 or more, and who passed the UKB’s internal quality check (i.e. “FILTER=PASS” in the vcf files). If a variant was found in both the genotype hard calls and the WES calls, the directly genotyped results were retained. As a last step to build this variant backbone, we submitted the resulting merged file for imputation with the TOPMed panel (October 6, 2022) and retained only variants with an imputation r^2^ > 0.95.

We then used a modified version of the HLA-TAPAS pipeline*34* to combine the HLA allele calls with the variant backbone above (“MakeReference”) and to impute HLA alleles in our cohort (“SNP2HLA”). No other parts of the HLA-TAPAS pipeline were used. Briefly, major modification to the HLA-TAPAS pipeline included the native usage of GRCh38 as the reference chromosome build (instead of successive lift-overs to hg19 and then to hg18), the use of Beagle v5.4 for phasing*47* and imputation*48*, the use of a “decoy” allele (99*99) to represent a lack of *HLA-DRB3*, *HLA-DRB4*, or *HLA-DRB5* alleles, and a longer list of genes which now includes *HLA-DMA*, *HLA-DMB*, *HLA-DOA*, *HLA-DOB*, *HLA-DRA*, *HLA-DRB3*, *HLA-DRB4*, *HLA-DRB5*, *HLA-E*, *HLA-F*, and *HLA-G*, on top of the previously available *HLA-A*, *HLA-B*, *HLA-C*, *HLA-DPA1*, *HLA-DPB1*, *HLA-DQA1*, *HLA-DQB1*, and *HLA-DRB1*. For the creation of the reference panel, we removed samples with a genotype missing rate of 30% or more, we removed genetic variants with a HWE p-value < 5×10^−8^ or a missing rate of 5% or more. For the SNP2HLA imputation step, imputed variables from our cohort were used as input (see ***Genotyping*** section above).

We did additional QC on the SNP2HLA results as follows. First, given that SNP2HLA assumes that HLA alleles are equivalent to biallelic single nucleotide variants, this can lead to situations where a participant will be assigned more than 2 alleles at any given genes. When this happened, we removed all imputed calls at that gene for that participant. Second, after comparing our imputed results with another HLA calling technology (see ***HLA imputation accuracy*** below), we selected an r^2^ threshold of 0.3 for all genes except for *HLA-DRB3*, *HLA-DRB4*, and *HLA-DRB5*, for which a threshold of 0.6 was used. Lastly, except for *HLA-DRB3*, *HLA-DRB4*, and *HLA-DRB5* which are not present in all individuals, genes for which an imputed call were available in less than 30% of participants were removed from any further analyses.

Lastly, once the final imputation calls were determined for each participant, we extracted their corresponding amino acid residue sequence using the IPD-IMGT/HLA database*49* (v3.45.0).

### HLA imputation accuracy

To assess the accuracy of our HLA imputation results, we directly genotyped 541 study participants using 454 sequencing technology (Roche, Basel, Switzerland) on the following genes: *HLA-A*, *HLA-B*, *HLA-DQA1*, *HLA-DQB1*, *HLA-DRB1*, *HLA-DRB3*, *HLA-DRB4*, and *HLA-DRB5*. 454 sequencing uses different combination of PCR primers for exon 2 to identify the HLA alleles. However, given that our imputation panel uses all exons to assign a HLA allele, this can lead to considerable ambiguity and falsely decrease the concordance between the two methods. Hence, all comparisons between the two methods were done at the G-group level. That is, we compared imputed and 454 sequencing called alleles at the level of exons 2 and 3 for class I genes, and exon 2 only for class II genes. Since these are the most polymorphic exons in a HLA gene, this is sufficient to assess how well the imputation panel performs*34*. Importantly, the concordance values obtained in this way are still also expected to be conservative estimates of the true performance of our imputation panel, since discordant results may still represent correctly imputed HLA alleles.

### GWAS and HLA association studies

For all association studies, serology levels were inverse qq-normalized to prevent variance inflation. All analyses used the first 10 genetic principal components, the cohort as a factor variable (CBC or PROVIDE), and participant sex as covariates. Age was not a covariate given that all participants were the same age on blood draw. To control for relatedness and polygenic risk, we used Regenie*32* for all GWAS and HLA allele and amino acid association studies. Regenie uses two steps. In step 1, a polygenic risk score is built to predict the phenotype. For this step, we only selected variants with a MAF > 1%, a minor allele count > 6, a genotype missing rate < 5%, HWE p-value > 10^−10^, and in linkage equilibrium (--indep-pairwise 200 100 0.2 in Plink v1.9). During step 2, Regenie uses this polygenic risk score with a leave-one-chromosome-out approach as additional covariates for the allele association studies. Only alleles with MAF > 0.5% were used for the GWAS, HLA allele, and amino acid associations studies.

For the haplotype association study for the HLA-DQ dimers, we could not use the usual GWAS additive effect model for single variants (i.e. a score of 0, 1, or 2 if the participant does not carry the allele, is heterozygous for the allele, or is homozygous for the allele, respectively). Instead, for any combination of *HLA-DQA1* and *HLA-DQB1* alleles, we gave a score of 0 if the participant did not have a pair of these alleles, a score of 1 if the participant is heterozygous for each allele, a score of 2 if the participant is homozygous for one allele and heterozygous for the other, and a score of 4 if the participant is homozygous for both alleles. This corresponds a dosage of allele pairs, mimicking the possible combination expected in an individual. This score was used as the regressor for the serology levels, and the linear regressions were performed in R v4.1.0*50*. The same covariates as above were used: sex, 10 genetic principal components, and Regenie’s step 1 polygenic risks.

We used a genome-wide significant threshold divided by the number of traits to determine statistical significance (i.e. 5×10^−8^/8). For the HLA allele association studies, we used the alleles that passed this threshold for conditional analyses. Conditional analyses were also performed with Regenie, with the same options and covariates as for the primary analyses above.

### HLA dimers visualization

HLA receptor dimers and amino acid residue association results were visualized with protein cartoon graphs using PyMOL*51*, then combined with the magick R package*52*. All dimer sequences and 3D structures were obtained from the Protein Data Bank*53*: 3LQZ*54* for HLA-DP, 7KEI for HLA-DQ, 4MDJ*55* for HLA-DRB1, and 1FV1*56* for HLA-DRB5.

### HLA receptor binding in-silico analyses

To determine HLA dimer binding to different HBsAg and DT epitopes, we first extracted their protein sequences from Uniprot*36* (January 20, 2023). The full search for HBsAG was “hbsag AND (virus_host_id9606)”, which was further narrowed only to Swiss-Prot reviewed sequences. The full search for DT was ‘(protein_name=“Diphtheria Toxin”) AND (length:[550 to 570])’. We then manually extracted sequences from any Corynebacterium organisms or from corynephage (*Diphtheria*’s viral bacteriophage which produces the toxin). As the number of available protein sequences was smaller for DT, we did not remove non-Swiss-Prot reviewed sequences. After removal of identical sequences, we were left with 50 HBsAg sequences and 13 DT sequences. We trimmed the beginning of each sequence to a length of 389 amino acids for HBsAg, and a length of 535 for DT, corresponding to the biologically active parts of each protein (pre-S and S for HBsAg*57*, and fragments A and B for DT*58*). Lastly, for DT, all analyses were done for fragment A and fragment B separately, as the full DT protein is cleaved to exert its full biological activity*58*.

We used NetMHCIIpan*35* (v4.0) to determine *in-silico* HLA dimer binding strength to all HBsAg and DT proteins obtained above. The default peptide length of 15 without context encoding was used. We used the eluted ligand results for all further analyses and their interpretation (i.e. the binding affinity mode was not used given its reduced accuracy*35*). For HLA-DP and HLA-DQ, analyses were restricted to dimers that were found in at least 5% of participants based on their HLA allele combination. For DRB dimers, analyses were restricted to corresponding *HLA-DRB1*, *HLA-DRB3*, *HLA-DRB4* and *HLA-DRB5* alleles with frequency > 10%. We used beta regression on the rank outputs (divided by 100) to compare relative *in-silico* binding between HLA dimers.

### Binding difference plots

To plot difference in *in-silico* binding strength between two HLA dimers, we subtracted the netMHCIIpan ranks of the worse dimer from those of the best dimer at each 15-mer peptide in the corresponding protein (HBsAg or DT). When there were multiple 15-mer for a given position, we used the one with the best binding strength for the subtraction. Hence the result from the subtraction is the difference between the best and the worst dimers for their “best case” binding strength at any given location of HBsAg.

### Ethics

For both Bangladeshi cohorts, written informed consents was obtained from either the mother only (PROVIDE) or either parents or guardians (CBC). For the PROVIDE cohort, the protocol and informed consent were approved by the Ethics Review Committee and the Research Review Committee of the International Centre for Diarrhoeal Disease Research, Bangladesh (ICDDR,B), the University of Virginia, and the University of Vermont. For the CBC cohort, the study was also approved by the same review boards at the ICDDR,B and the University of Virginia. The genetic component of the study was also independently approved by the Oxford Tropical Research Ethics Committee.

For the replication cohorts from Africa studied as part of VaccGene, ethical approval was provided locally by the Uganda Virus Research Institute (reference GC/127/12/07/32) and Uganda National Council for Science and Technology (MV625), and in the UK by London School of Hygiene and Tropical Medicine (A340) and Oxford Tropical Research (39-12 and 42-14) Ethics Committees for the Uganda cohort. Ethical approval was obtained from ethical approval from the University of Witwatersrand and Human Research Ethics Committee (reference M130714) and the Oxford Tropical Research Ethics Committee (1042-13 and 42-14) for the South African cohort. Lastly, ethical approval was obtained from the Ministere de la Recherche Scientifique et de l’Innovation in Burkina Faso (reference 2014-12-151) and the Oxford Tropical Research Ethics Committee (41-12).

For the UK Biobank, ethical approval was obtained from the North West Multi-centre Research Ethics Committee (https://www.hra.nhs.uk/about-us/committees-and-services/res-and-recs/search-research-ethics-committees/north-west-haydock/).

## Data Availability

All direct genotypes from the Bangladeshi individuals post-quality control alongside imputed data and raw and curated HLA sequence data and calls have been submitted to the European Genome-Phenome Archive under accession EGAS00001000918. Summary statistics for the genome-wide association tests of imputed data for eight vaccine antibody levels are available on the GWAS Catalog. Code to the modified HLA-TAPAS/SNP2HLA software is available on github at https://github.com/DrGBL/snp2hla_redux.

## Acknowledgements

We thank all sample donors who contributed to this study and staff involved in consenting, sample and data collection and preparation including interviewers, computer and laboratory technicians, clerical workers, research scientists, volunteers, managers, receptionists, and nurses. We are grateful to Steve Rich and Joe Mychaleckyj for assistance with elements of the genome-wide association study. This research has been conducted using the UK Biobank Resource under Application Number 27449.

## Author contributions

Conceptualization: GBL, AJM

Data curation: GBL, KA, TC, CP

Formal Analysis: GBL, AJM

Funding acquisition: GBL, JBR, AVSH, RH, WAP, AJM

Investigation: GBL, GLW, MK, MA, MSS, RH

Methodology: GBL, AVSH, PD, MSS, RH, WAP, AJM

Project administration: AVSH, PD, MDD, RH, WAP, AJM

Resources: JAN, GS, SW, FRMvdK, KJ, JBR, AVSH, PD, MSS, RH, WAP, AJM

Software: GBL, AYC

Supervision: AJM

Validation: GBL, JAN, SLM, BDK, SS, SM, AE

Visualization: GBL, AJM

Writing – original draft: GBL, AJM

Writing – review & editing: GBL, AYC, JAN, BK, JBR, PD, RH WAP, AJM

## Supplementary Materials

Materials and Methods

Supplementary Text

Figs. S1 to S11

Tables S1 to S3

Data S1 to S7

## References and notes

1. M. G. Hossain, K. Ueda, A meta-analysis on genetic variability of RT/HBsAg overlapping region of hepatitis B virus (HBV) isolates of Bangladesh. Infect. Agent. Cancer. 14, 33 (2019).

2. J. Cremer, S. H. I. Hofstraat, F. van Heiningen, I. K. Veldhuijzen, B. H. B. van Benthem, K. S. M. Benschop, Genetic variation of hepatitis B surface antigen among acute and chronic hepatitis B virus infections in The Netherlands. J. Med. Virol. 90, 1576–1585 (2018).

3. S. U. Munshi, T. T. T. Tran, T. N. T. Vo, S. Tabassum, N. Sultana, T. H. Nguyen, M. Jahan, C. N. Le, S. Baker, M. Rahman, Molecular characterization of hepatitis B virus in Bangladesh reveals a highly recombinant population. PLoS One. 12, e0188944 (2017).

4. T. Inoue, Y. Tanaka, Cross-Protection of Hepatitis B Vaccination among Different Genotypes. Vaccines. 8 (2020), doi:10.3390/vaccines8030456.

5. M.-H. Chang, Breakthrough HBV infection in vaccinated children in Taiwan: surveillance for HBV mutants. Antivir. Ther. 15, 463–469 (2010).

6. H.-Y. Hsu, M.-H. Chang, S.-H. Liaw, Y.-H. Ni, H.-L. Chen, Changes of hepatitis B surface antigen variants in carrier children before and after universal vaccination in taiwan. Hepatology. 30, 1312–1317 (1999).

7. J. N. Zuckerman, Protective efficacy, immunotherapeutic potential, and safety of hepatitis B vaccines. J. Med. Virol. 78, 169–177 (2006).

8. W. J. McAleer, E. B. Buynak, R. Z. Maigetter, D. E. Wampler, W. J. Miller, M. R. Hilleman, Human hepatitis B vaccine from recombinant yeast. Nature. 307, 178–180 (1984).

9. European Medicines Agency, European Medicines Agency, (available at https://www.ema.europa.eu/en/medicines).

10. Centers for Disease Control and Prevention, Hepatitis B Information, (available at https://www.cdc.gov/hepatitis/hbv/hbvfaq.htm).

11. UK Health Security Agency, UK Immunisation, (available at https://www.gov.uk/government/collections/immunisation).

12. P. Rendi-Wagner, D. Shouval, B. Genton, Y. Lurie, H. Rümke, G. Boland, A. Cerny, M. Heim, D. Bach, M. Schroeder, H. Kollaritsch, Comparative immunogenicity of a PreS/S hepatitis B vaccine in non- and low responders to conventional vaccine. Vaccine. 24, 2781–2789 (2006).

13. J. S. J. Bertino, P. Tirrell, R. N. Greenberg, H. L. Keyserling, G. A. Poland, D. Gump, M. L. Kumar, K. Ramsey, A comparative trial of standard or high-dose S subunit recombinant hepatitis B vaccine versus a vaccine containing S subunit, pre-S1, and pre-S2 particles for revaccination of healthy adult nonresponders. J. Infect. Dis. 175, 678–681 (1997).

14. A. Hourvitz, R. Mosseri, A. Solomon, Y. Yehezkelli, J. Atsmon, Y. L. Danon, R. Koren, D. Shouval, Reactogenicity and immunogenicity of a new recombinant hepatitis B vaccine containing Pre S antigens: a preliminary report. J. Viral Hepat. 3, 37–42 (1996).

15. J. N. Zuckerman, C. Sabin, F. M. Craig, A. Williams, A. J. Zuckerman, Immune response to a new hepatitis B vaccine in healthcare workers who had not responded to standard vaccine: randomised double blind dose-response study. BMJ. 314, 329–333 (1997).

16. E. V Esaulenko, A. A. Yakovlev, G. A. Volkov, A. A. Sukhoruk, K. G. Surkov, P. V Kruglyakov, F. Diaz-Mitoma, Efficacy and Safety of a 3-Antigen (Pre-S1/Pre-S2/S) Hepatitis B Vaccine: Results of a Phase 3 Randomized Clinical Trial in the Russian Federation. Clin. Infect. Dis. an Off. Publ. Infect. Dis. Soc. Am. 73, e3333–e3339 (2021).

17. T. Vesikari, A. Finn, P. van Damme, I. Leroux-Roels, G. Leroux-Roels, N. Segall, A. Toma, G. Vallieres, R. Aronson, D. Reich, S. Arora, P. J. Ruane, C. L. Cone, M. Manns, C. Cosgrove, S. N. Faust, M. N. Ramasamy, N. Machluf, J. N. Spaans, B. Yassin-Rajkumar, D. Anderson, V. Popovic, F. Diaz-Mitoma, Immunogenicity and Safety of a 3-Antigen Hepatitis B Vaccine vs a Single-Antigen Hepatitis B Vaccine: A Phase 3 Randomized Clinical Trial. JAMA Netw. open. 4, e2128652 (2021).

18. A. J. Mentzer, D. O’Connor, S. Bibi, I. Chelysheva, E. A. Clutterbuck, T. Demissie, T. Dinesh, N. J. Edwards, S. Felle, S. Feng, A. L. Flaxman, E. Karp-Tatham, G. Li, X. Liu, N. Marchevsky, L. Godfrey, R. Makinson, M. B. Bull, J. Fowler, B. Alamad, T. Malinauskas, A. Y. Chong, K. Sanders, R. H. Shaw, M. Voysey, M. D. Snape, A. J. Pollard, T. Lambe, J. C. Knight, Human leukocyte antigen alleles associate with COVID-19 vaccine immunogenicity and risk of breakthrough infection. Nat. Med. 29, 147–157 (2023).

19. J. Bovijn, C. M. Lindgren, M. V Holmes, Genetic variants mimicking therapeutic inhibition of IL-6 receptor signaling and risk of COVID-19. Lancet Rheumatol. 2, e658– e659 (2020).

20. RECOVERY Collaborative Group, Tocilizumab in patients admitted to hospital with COVID-19 (RECOVERY): a randomised, controlled, open-label, platform trial. Lancet (London, England). 397, 1637–1645 (2021).

21. G. Butler-Laporte, T. Nakanishi, V. Mooser, D. R. Morrison, T. Abdullah, O. Adeleye, N. Mamlouk, N. Kimchi, Z. Afrasiabi, N. Rezk, A. Giliberti, A. Renieri, Y. Chen, S. Zhou, V. Forgetta, J. B. Richards, Vitamin D and COVID-19 susceptibility and severity in the COVID-19 Host Genetics Initiative: A Mendelian randomization study. PLOS Med. 18, e1003605 (2021).

22. I. H. Murai, A. L. Fernandes, L. P. Sales, A. J. Pinto, K. F. Goessler, C. S. C. Duran, C. B. R. Silva, A. S. Franco, M. B. Macedo, H. H. H. Dalmolin, J. Baggio, G. G. M. Balbi, B. Z. Reis, L. Antonangelo, V. F. Caparbo, B. Gualano, R. M. R. Pereira, Effect of a Single High Dose of Vitamin D3 on Hospital Length of Stay in Patients With Moderate to Severe COVID-19: A Randomized Clinical Trial. JAMA (2021), doi:10.1001/jama.2020.26848.

23. G. L. Wojcik, C. Marie, M. M. Abhyankar, N. Yoshida, K. Watanabe, A. J. Mentzer, T. Carstensen, J. Mychaleckyj, B. D. Kirkpatrick, S. S. Rich, P. Concannon, R. Haque, G. C. Tsokos, W. A. J. Petri, P. Duggal, Genome-Wide Association Study Reveals Genetic Link between Diarrhea-Associated Entamoeba histolytica Infection and Inflammatory Bowel Disease. MBio. 9 (2018), doi:10.1128/mBio.01668-18.

24. G. L. Wojcik, P. Korpe, C. Marie, A. J. Mentzer, T. Carstensen, J. Mychaleckyj, B. D. Kirkpatrick, S. S. Rich, P. Concannon, A. S. G. Faruque, R. Haque, W. A. J. Petri, P. Duggal, Genome-Wide Association Study of Cryptosporidiosis in Infants Implicates PRKCA. MBio. 11 (2020), doi:10.1128/mBio.03343-19.

25. B. D. Kirkpatrick, E. R. Colgate, J. C. Mychaleckyj, R. Haque, D. M. Dickson, M. P. Carmolli, U. Nayak, M. Taniuchi, C. Naylor, F. Qadri, J. Z. Ma, M. Alam, M. C. Walsh, S. A. Diehl, W. A. J. Petri, The “Performance of Rotavirus and Oral Polio Vaccines in Developing Countries” (PROVIDE) study: description of methods of an interventional study designed to explore complex biologic problems. Am. J. Trop. Med. Hyg. 92, 744– 751 (2015).

26. C. A. Gilchrist, J. A. Cotton, C. Burkey, T. Arju, A. Gilmartin, Y. Lin, E. Ahmed, K. Steiner, M. Alam, S. Ahmed, G. Robinson, S. U. Zaman, M. Kabir, M. Sanders, R. M. Chalmers, T. Ahmed, J. Z. Ma, R. Haque, A. S. G. Faruque, M. Berriman, W. A. Petri, Genetic Diversity of Cryptosporidium hominis in a Bangladeshi Community as Revealed by Whole-Genome Sequencing. J. Infect. Dis. 218, 259–264 (2018).

27. D. Duchen, R. Haque, L. Chen, G. Wojcik, P. Korpe, U. Nayak, A. J. Mentzer, B. Kirkpatrick, W. A. J. Petri, P. Duggal, Host Genome-Wide Association Study of Infant Susceptibility to Shigella-Associated Diarrhea. Infect. Immun. 89 (2021), doi:10.1128/IAI.00012-21.

28. R. M. Munday, R. Haque, N.-J. Jan, G. L. Wojcik, C. Marie, D. Duchen, A. J. Mentzer, U. Nayak, P. Korpe, B. D. Kirkpatrick, W. A. J. Petri, P. Duggal, Genome-Wide Association Study of Campylobacter-Positive Diarrhea Identifies Genes Involved in Toxin Processing and Inflammatory Response. MBio. 13, e0055622 (2022).

29. R. M. Munday, R. Haque, G. L. Wojcik, P. Korpe, U. Nayak, B. D. Kirkpatrick, W. A. Petri, P. Duggal, Genome-wide association studies of diarrhea frequency and duration in the first year of life in Bangladeshi infants. J. Infect. Dis. (2023), doi:10.1093/infdis/jiad068.

30. K. Estrada, U. Styrkarsdottir, E. Evangelou, Y.-H. Hsu, E. L. Duncan, E. E. Ntzani, L. Oei, O. M. E. Albagha, N. Amin, J. P. Kemp, D. L. Koller, G. Li, C.-T. Liu, R. L. Minster, A. Moayyeri, L. Vandenput, D. Willner, S.-M. Xiao, L. M. Yerges-Armstrong, H.-F. Zheng, N. Alonso, J. Eriksson, C. M. Kammerer, S. K. Kaptoge, P. J. Leo, G. Thorleifsson, S. G. Wilson, J. F. Wilson, V. Aalto, M. Alen, A. K. Aragaki, T. Aspelund, J. R. Center, Z. Dailiana, D. J. Duggan, M. Garcia, N. Garcia-Giralt, S. Giroux, G. Hallmans, L. J. Hocking, L. B. Husted, K. A. Jameson, R. Khusainova, G. S. Kim, C. Kooperberg, T. Koromila, M. Kruk, M. Laaksonen, A. Z. Lacroix, S. H. Lee, P. C. Leung, J. R. Lewis, L. Masi, S. Mencej-Bedrac, T. V Nguyen, X. Nogues, M. S. Patel, J. Prezelj, L. M. Rose, S. Scollen, K. Siggeirsdottir, A. V Smith, O. Svensson, S. Trompet, O. Trummer, N. M. van Schoor, J. Woo, K. Zhu, S. Balcells, M. L. Brandi, B. M. Buckley, S. Cheng, C. Christiansen, C. Cooper, G. Dedoussis, I. Ford, M. Frost, D. Goltzman, J. González-Macías, M. Kähönen, M. Karlsson, E. Khusnutdinova, J.-M. Koh, P. Kollia, B. L. Langdahl, W. D. Leslie, P. Lips, Ö. Ljunggren, R. S. Lorenc, J. Marc, D. Mellström, B. Obermayer-Pietsch, J. M. Olmos, U. Pettersson-Kymmer, D. M. Reid, J. A. Riancho, P. M. Ridker, F. Rousseau, P. E. S. Lagboom, N. L. S. Tang, R. Urreizti, W. Van Hul, J. Viikari, M. T. Zarrabeitia, Y. S. Aulchenko, M. Castano-Betancourt, E. Grundberg, L. Herrera, T. Ingvarsson, H. Johannsdottir, T. Kwan, R. Li, R. Luben, C. Medina-Gómez, S. Th Palsson, S. Reppe, J. I. Rotter, G. Sigurdsson, J. B. J. van Meurs, D. Verlaan, F. M. K. Williams, A. R. Wood, Y. Zhou, K. M. Gautvik, T. Pastinen, S. Raychaudhuri, J. A. Cauley, D. I. Chasman, G. R. Clark, S. R. Cummings, P. Danoy, E. M. Dennison, R. Eastell, J. A. Eisman, V. Gudnason, A. Hofman, R. D. Jackson, G. Jones, J. W. Jukema, K.-T. Khaw, T. Lehtimäki, Y. Liu, M. Lorentzon, E. McCloskey, B. D. Mitchell, K. Nandakumar, G. C. Nicholson, B. A. Oostra, M. Peacock, H. A. P. Pols, R. L. Prince, O. Raitakari, I. R. Reid, J. Robbins, P. N. Sambrook, P. C. Sham, A. R. Shuldiner, F. A. Tylavsky, C. M. van Duijn, N. J. Wareham, L. A. Cupples, M. J. Econs, D. M. Evans, T. Harris, A. W. C. Kung, B. M. Psaty, J. Reeve, T. D. Spector, E. A. Streeten, M. C. Zillikens, U. Thorsteinsdottir, C. Ohlsson, D. Karasik, J. B. Richards, M. A. Brown, K. Stefansson, A. G. Uitterlinden, S. H. Ralston, J. P. A. Ioannidis, D. P. Kiel, F. Rivadeneira, Genome-wide meta-analysis identifies 56 bone mineral density loci and reveals 14 loci associated with risk of fracture. Nat. Genet. 44, 491–501 (2012).

31. D. Taliun, D. N. Harris, M. D. Kessler, J. Carlson, Z. A. Szpiech, R. Torres, S. A. G. Taliun, A. Corvelo, S. M. Gogarten, H. M. Kang, A. N. Pitsillides, J. LeFaive, S. Lee, X. Tian, B. L. Browning, S. Das, A.-K. Emde, W. E. Clarke, D. P. Loesch, A. C. Shetty, T. W. Blackwell, A. V Smith, Q. Wong, X. Liu, M. P. Conomos, D. M. Bobo, F. Aguet, C. Albert, A. Alonso, K. G. Ardlie, D. E. Arking, S. Aslibekyan, P. L. Auer, J. Barnard, R. G. Barr, L. Barwick, L. C. Becker, R. L. Beer, E. J. Benjamin, L. F. Bielak, J. Blangero, M. Boehnke, D. W. Bowden, J. A. Brody, E. G. Burchard, B. E. Cade, J. F. Casella, B. Chalazan, D. I. Chasman, Y.-D. I. Chen, M. H. Cho, S. H. Choi, M. K. Chung, C. B. Clish, A. Correa, J. E. Curran, B. Custer, D. Darbar, M. Daya, M. de Andrade, D. L. DeMeo, S. K. Dutcher, P. T. Ellinor, L. S. Emery, C. Eng, D. Fatkin, T. Fingerlin, L. Forer, M. Fornage, N. Franceschini, C. Fuchsberger, S. M. Fullerton, S. Germer, M. T. Gladwin, D. J. Gottlieb, X. Guo, M. E. Hall, J. He, N. L. Heard-Costa, S. R. Heckbert, M. R. Irvin, J. M. Johnsen, A. D. Johnson, R. Kaplan, S. L. R. Kardia, T. Kelly, S. Kelly, E. E. Kenny, D. P. Kiel, R. Klemmer, B. A. Konkle, C. Kooperberg, A. Köttgen, L. A. Lange, J. Lasky-Su, D. Levy, X. Lin, K.-H. Lin, C. Liu, R. J. F. Loos, L. Garman, R. Gerszten, S. A. Lubitz, K. L. Lunetta, A. C. Y. Mak, A. Manichaikul, A. K. Manning, R. A. Mathias, D. D. McManus, S. T. McGarvey, J. B. Meigs, D. A. Meyers, J. L. Mikulla, M. A. Minear, B. D. Mitchell, S. Mohanty, M. E. Montasser, C. Montgomery, A. C. Morrison, J. M. Murabito, A. Natale, P. Natarajan, S. C. Nelson, K. E. North, J. R. O’Connell, N. D. Palmer, N. Pankratz, G. M. Peloso, P. A. Peyser, J. Pleiness, W. S. Post, B. M. Psaty, D. C. Rao, S. Redline, A. P. Reiner, D. Roden, J. I. Rotter, I. Ruczinski, C. Sarnowski, S. Schoenherr, D. A. Schwartz, J.-S. Seo, S. Seshadri, V. A. Sheehan, W. H. Sheu, M. B. Shoemaker, N. L. Smith, J. A. Smith, N. Sotoodehnia, A. M. Stilp, W. Tang, K. D. Taylor, M. Telen, T. A. Thornton, R. P. Tracy, D. J. Van Den Berg, R. S. Vasan, K. A. Viaud-Martinez, S. Vrieze, D. E. Weeks, B. S. Weir, S. T. Weiss, L.-C. Weng, C. J. Willer, Y. Zhang, X. Zhao, D. K. Arnett, A. E. Ashley-Koch, K. C. Barnes, E. Boerwinkle, S. Gabriel, R. Gibbs, K. M. Rice, S. S. Rich, E. K. Silverman, P. Qasba, W. Gan, N. Abe, L. Almasy, S. Ament, P. Anderson, P. Anugu, D. Applebaum-Bowden, T. Assimes, D. Avramopoulos, E. Barron-Casella, T. Beaty, G. Beck, D. Becker, A. Beitelshees, T. Benos, M. Bezerra, J. Bis, R. Bowler, U. Broeckel, J. Broome, K. Bunting, C. Bustamante, E. Buth, J. Cardwell, V. Carey, C. Carty, R. Casaburi, P. Castaldi, M. Chaffin, C. Chang, Y.-C. Chang, S. Chavan, B.-J. Chen, W.-M. Chen, L.-M. Chuang, R.-H. Chung, S. Comhair, E. Cornell, C. Crandall, J. Crapo, J. Curtis, C. Damcott, S. David, C. Davis, L. de las Fuentes, M. DeBaun, R. Deka, S. Devine, Q. Duan, R. Duggirala, J. P. Durda, C. Eaton, L. Ekunwe, A. El Boueiz, S. Erzurum, C. Farber, M. Flickinger, M. Fornage, C. Frazar, M. Fu, L. Fulton, S. Gao, Y. Gao, M. Gass, B. Gelb, X. P. Geng, M. Geraci, A. Ghosh, C. Gignoux, D. Glahn, D.-W. Gong, H. Goring, S. Graw, D. Grine, C. C. Gu, Y. Guan, N. Gupta, J. Haessler, N. L. Hawley, B. Heavner, D. Herrington, C. Hersh, B. Hidalgo, J. Hixson, B. Hobbs, J. Hokanson, E. Hong, K. Hoth, C. A. Hsiung, Y.-J. Hung, H. Huston, C. M. Hwu, R. Jackson, D. Jain, M. A. Jhun, C. Johnson, R. Johnston, K. Jones, S. Kathiresan, A. Khan, W. Kim, G. Kinney, H. Kramer, C. Lange, E. Lange, L. Lange, C. Laurie, M. LeBoff, J. Lee, S. S. Lee, W.-J. Lee, D. Levine, J. Lewis, X. Li, Y. Li, H. Lin, H. Lin, K. H. Lin, S. Liu, Y. Liu, Y. Liu, J. Luo, M. Mahaney, N. T.- O. for P. M. (TOPMed) Consortium, Sequencing of 53,831 diverse genomes from the NHLBI TOPMed Program. Nature. 590, 290–299 (2021).

32. J. Mbatchou, L. Barnard, J. Backman, A. Marcketta, J. A. Kosmicki, A. Ziyatdinov, C. Benner, C. O’Dushlaine, M. Barber, B. Boutkov, L. Habegger, M. Ferreira, A. Baras, J. Reid, G. Abecasis, E. Maxwell, J. Marchini, Computationally efficient whole-genome regression for quantitative and binary traits. Nat. Genet. 53, 1097–1103 (2021).

33. G. Butler-Laporte, J. Farjoun, T. Nakanishi, Y. Chen, M. Hultström, T. Lu, S. Yoshiji, Y. Ilboudo, K. Y. H. Liang, C.-Y. Su, J. D. S. Willett, S. Zhou, V. Forgetta, D. Taliun, J. B. Richards, medRxiv, in press, doi:10.1101/2023.01.15.23284570.

34. Y. Luo, M. Kanai, W. Choi, X. Li, S. Sakaue, K. Yamamoto, K. Ogawa, M. Gutierrez-Arcelus, P. K. Gregersen, P. E. Stuart, J. T. Elder, L. Forer, S. Schönherr, C. Fuchsberger, A. V Smith, J. Fellay, M. Carrington, D. W. Haas, X. Guo, N. D. Palmer, Y.-D. I. Chen, J. I. Rotter, K. D. Taylor, S. S. Rich, A. Correa, J. G. Wilson, S. Kathiresan, M. H. Cho, A. Metspalu, T. Esko, Y. Okada, B. Han, N. Abe, G. Abecasis, F. Aguet, C. Albert, L. Almasy, A. Alonso, S. Ament, P. Anderson, P. Anugu, D. Applebaum-Bowden, K. Ardlie, D. Arking, D. K. Arnett, A. Ashley-Koch, S. Aslibekyan, T. Assimes, P. Auer, D. Avramopoulos, N. Ayas, A. Balasubramanian, J. Barnard, K. Barnes, R. G. Barr, E. Barron-Casella, L. Barwick, T. Beaty, G. Beck, D. Becker, L. Becker, R. Beer, A. Beitelshees, E. Benjamin, T. Benos, M. Bezerra, L. Bielak, J. Bis, T. Blackwell, J. Blangero, E. Boerwinkle, D. W. Bowden, R. Bowler, J. Brody, U. Broeckel, J. Broome, D. Brown, K. Bunting, E. Burchard, C. Bustamante, E. Buth, B. Cade, J. Cardwell, V. Carey, J. Carrier, C. Carty, R. Casaburi, J. P. C. Romero, J. Casella, P. Castaldi, M. Chaffin, C. Chang, Y.-C. Chang, D. Chasman, S. Chavan, B.-J. Chen, W.-M. Chen, S. H. Choi, L.-M. Chuang, M. Chung, R.-H. Chung, C. Clish, S. Comhair, M. Conomos, E. Cornell, C. Crandall, J. Crapo, L. A. Cupples, J. Curran, J. Curtis, B. Custer, C. Damcott, D. Darbar, S. David, C. Davis, M. Daya, M. de Andrade, L. de las Fuentes, P. de Vries, M. DeBaun, R. Deka, D. DeMeo, S. Devine, H. Dinh, H. Doddapaneni, Q. Duan, S. Dugan-Perez, R. Duggirala, J. P. Durda, S. K. Dutcher, C. Eaton, L. Ekunwe, A. El Boueiz, P. Ellinor, L. Emery, S. Erzurum, C. Farber, J. Farek, T. Fingerlin, M. Flickinger, M. Fornage, N. Franceschini, C. Frazar, M. Fu, S. M. Fullerton, L. Fulton, S. Gabriel, W. Gan, S. Gao, Y. Gao, M. Gass, H. Geiger, B. Gelb, M. Geraci, S. Germer, R. Gerszten, A. Ghosh, R. Gibbs, C. Gignoux, M. Gladwin, D. Glahn, S. Gogarten, D.-W. Gong, H. Goring, S. Graw, K. J. Gray, D. Grine, C. Gross, C. C. Gu, Y. Guan, N. Gupta, D. M. Haas, J. Haessler, M. Hall, Y. Han, P. Hanly, D. Harris, N. L. Hawley, J. He, B. Heavner, S. Heckbert, R. Hernandez, D. Herrington, C. Hersh, B. Hidalgo, J. Hixson, B. Hobbs, J. Hokanson, E. Hong, K. Hoth, C. (Agnes) Hsiung, J. Hu, Y.-J. Hung, H. Huston, C. M. Hwu, M. R. Irvin, R. Jackson, D. Jain, C. Jaquish, J. Johnsen, A. Johnson, C. Johnson, R. Johnston, K. Jones, H. M. Kang, R. Kaplan, S. Kardia, S. Kelly, E. Kenny, M. Kessler, A. Khan, Z. Khan, W. Kim, J. Kimoff, G. Kinney, B. Konkle, C. Kooperberg, H. Kramer, C. Lange, E. Lange, L. Lange, C. Laurie, C. Laurie, M. LeBoff, J. Lee, S. Lee, W.-J. Lee, J. LeFaive, D. Levine, D. Levy, J. Lewis, X. Li, Y. Li, H. Lin, H. Lin, X. Lin, S. Liu, Y. Liu, Y. Liu, R. J. F. Loos, S. Lubitz, K. Lunetta, J. Luo, U. Magalang, M. Mahaney, B. Make, A. Manichaikul, A. Manning, J. Manson, L. Martin, M. Marton, S. Mathai, R. Mathias, S. May, P. McArdle, M.-L. McDonald, S. McFarland, S. McGarvey, D. McGoldrick, C. McHugh, B. McNeil, H. Mei, J. Meigs, V. Menon, L. Mestroni, G. Metcalf, D. A. Meyers, E. Mignot, J. Mikulla, N. Min, M. Minear, R. L. Minster, B. D. Mitchell, M. Moll, Z. Momin, M. E. Montasser, C. Montgomery, D. Muzny, J. C. Mychaleckyj, G. Nadkarni, R. Naik, T. Naseri, P. Natarajan, S. Nekhai, S. C. Nelson, B. Neltner, C. Nessner, D. Nickerson, O. Nkechinyere, K. North, J. O’Connell, T. O’Connor, H. Ochs-Balcom, G. Okwuonu, A. Pack, D. T. Paik, J. Pankow, G. Papanicolaou, C. Parker, N. T.-O. for P. M. (TOPMed) Consortium, A high-resolution HLA reference panel capturing global population diversity enables multi-ancestry fine-mapping in HIV host response. Nat. Genet. 53, 1504–1516 (2021).

35. B. Reynisson, C. Barra, S. Kaabinejadian, W. H. Hildebrand, B. Peters, M. Nielsen, Improved Prediction of MHC II Antigen Presentation through Integration and Motif Deconvolution of Mass Spectrometry MHC Eluted Ligand Data. J. Proteome Res. 19, 2304–2315 (2020).

36. UniProt: the Universal Protein Knowledgebase in 2023. Nucleic Acids Res. 51, D523–D531 (2023).

37. A. J. Mentzer, A. T. Dilthey, M. Pollard, D. Gurdasani, E. Karakoc, T. Carstensen, A. Muhwezi, C. Cutland, A. Diarra, R. da S. Antunes, S. Paul, G. Smits, S. Wareing, H. Kim, C. Pomilla, A. Y. Chong, D. Y. C. Brandt, R. Nielsen, S. Neaves, N. Timpson, A. Crinklaw, C. S. Lindestam Arlehamn, A. Rautanen, D. Kizito, T. Parks, K. Auckland, K. E. Elliott, T. Mills, K. Ewer, N. Edwards, S. Fatumo, S. Peacock, K. Jeffery, F. R. M. van der Klis, P. Kaleebu, P. Vijayanand, B. Peters, A. Sette, N. Cereb, S. Sirima, S. A. Madhi, A. M. Elliott, G. McVean, A. V. S. Hill, M. S. Sandhu, medRxiv, in press, doi:10.1101/2022.11.24.22282715.

38. B. Yan, J. Lv, Y. Feng, J. Liu, F. Ji, A. Xu, L. Zhang, Temporal trend of hepatitis B surface mutations in the post-immunization period: 9 years of surveillance (2005-2013) in eastern China. Sci. Rep. 7, 6669 (2017).

39. E. Tabor, Infections by hepatitis B surface antigen gene mutants in Europe and North America. J. Med. Virol. 78 Suppl 1, S43–7 (2006).

40. P. G. M. van Gageldonk, F. G. van Schaijk, F. R. van der Klis, G. A. M. Berbers, Development and validation of a multiplex immunoassay for the simultaneous determination of serum antibodies to Bordetella pertussis, diphtheria and tetanus. J. Immunol. Methods. 335, 79–89 (2008).

41. G. P. Smits, P. G. van Gageldonk, L. M. Schouls, F. R. M. van der Klis, G. A. M. Berbers, Development of a bead-based multiplex immunoassay for simultaneous quantitative detection of IgG serum antibodies against measles, mumps, rubella, and varicella-zoster virus. Clin. Vaccine Immunol. 19, 396–400 (2012).

42. R. M. de Voer, R. M. Schepp, F. G. A. Versteegh, F. R. M. van der Klis, G. A. M. Berbers, Simultaneous detection of Haemophilus influenzae type b polysaccharide-specific antibodies and Neisseria meningitidis serogroup A, C, Y, and W-135 polysaccharide-specific antibodies in a fluorescent-bead-based multiplex immunoassay. Clin. Vaccine Immunol. 16, 433–436 (2009).

43. I. D. Brinkman, J. de Wit, G. P. Smits, H. I. Ten Hulscher, M. C. Jongerius, T. C. Abreu, F. R. M. van der Klis, S. J. M. Hahné, M. P. G. Koopmans, N. Y. Rots, D. van Baarle, R. S. van Binnendijk, Early Measles Vaccination During an Outbreak in the Netherlands: Short-Term and Long-Term Decreases in Antibody Responses Among Children Vaccinated Before 12 Months of Age. J. Infect. Dis. 220, 594–602 (2019).

44. E. M. Swart, P. G. M. van Gageldonk, H. E. de Melker, F. R. van der Klis, G. A. M. Berbers, L. Mollema, Long-Term Protection against Diphtheria in the Netherlands after 50 Years of Vaccination: Results from a Seroepidemiological Study. PLoS One. 11, e0148605 (2016).

45. C. Bycroft, C. Freeman, D. Petkova, G. Band, L. T. Elliott, K. Sharp, A. Motyer, D. Vukcevic, O. Delaneau, J. O’Connell, A. Cortes, S. Welsh, A. Young, M. Effingham, G. McVean, S. Leslie, N. Allen, P. Donnelly, J. Marchini, The UK Biobank resource with deep phenotyping and genomic data. Nature. 562, 203–209 (2018).

46. J. D. Backman, A. H. Li, A. Marcketta, D. Sun, J. Mbatchou, M. D. Kessler, C. Benner, D. Liu, A. E. Locke, S. Balasubramanian, A. Yadav, N. Banerjee, C. E. Gillies, A. Damask, S. Liu, X. Bai, A. Hawes, E. Maxwell, L. Gurski, K. Watanabe, J. A. Kosmicki, V. Rajagopal, J. Mighty, M. Jones, L. Mitnaul, E. Stahl, G. Coppola, E. Jorgenson, L. Habegger, W. J. Salerno, A. R. Shuldiner, L. A. Lotta, J. D. Overton, M. N. Cantor, J. G. Reid, G. Yancopoulos, H. M. Kang, J. Marchini, A. Baras, G. R. Abecasis, M. A. R. Ferreira, R. G. Center, DiscovEHR, Exome sequencing and analysis of 454,787 UK Biobank participants. Nature. 599, 628–634 (2021).

47. B. L. Browning, X. Tian, Y. Zhou, S. R. Browning, Fast two-stage phasing of large-scale sequence data. Am. J. Hum. Genet. 108, 1880–1890 (2021).

48. B. L. Browning, Y. Zhou, S. R. Browning, A One-Penny Imputed Genome from Next-Generation Reference Panels. Am. J. Hum. Genet. 103, 338–348 (2018).

49. D. J. Barker, G. Maccari, X. Georgiou, M. A. Cooper, P. Flicek, J. Robinson, S. G. E. Marsh, The IPD-IMGT/HLA Database. Nucleic Acids Res. 51, D1053–D1060 (2023).

50. R Core Team, R: A Language and Environment for Statistical Computing (2021).

51. Schrödinger LLC., The PyMOL Molecular Graphics System, Version 2.5.4.

52. J. Ooms, magick: Advanced Graphics and Image-Processing in R (2021), (available at https://cran.r-project.org/package=magick).

53. H. M. Berman, J. Westbrook, Z. Feng, G. Gilliland, T. N. Bhat, H. Weissig, I. N. Shindyalov, P. E. Bourne, The Protein Data Bank. Nucleic Acids Res. 28, 235–242 (2000).

54. S. Dai, G. A. Murphy, F. Crawford, D. G. Mack, M. T. Falta, P. Marrack, J. W. Kappler, A. P. Fontenot, Crystal structure of HLA-DP2 and implications for chronic beryllium disease. Proc. Natl. Acad. Sci. U. S. A. 107, 7425–7430 (2010).

55. S. W. Scally, J. Petersen, S. C. Law, N. L. Dudek, H. J. Nel, K. L. Loh, L. C. Wijeyewickrema, S. B. G. Eckle, J. van Heemst, R. N. Pike, J. McCluskey, R. E. Toes, N. L. La Gruta, A. W. Purcell, H. H. Reid, R. Thomas, J. Rossjohn, A molecular basis for the association of the HLA-DRB1 locus, citrullination, and rheumatoid arthritis. J. Exp. Med. 210, 2569–2582 (2013).

56. Y. Li, H. Li, R. Martin, R. A. Mariuzza, Structural basis for the binding of an immunodominant peptide from myelin basic protein in different registers by two HLA-DR2 proteins. J. Mol. Biol. 304, 177–188 (2000).

57. S. Urban, R. Bartenschlager, R. Kubitz, F. Zoulim, Strategies to inhibit entry of HBV and HDV into hepatocytes. Gastroenterology. 147, 48–64 (2014).

58. S. F. Carroll, J. T. Barbieri, R. B. T.-M. in E. John Collier, “[11] Diphtheria toxin: Purification and properties” in Microbial Toxins: Tools in Enzymology (Academic Press, 1988; https://www.sciencedirect.com/science/article/pii/S0076687988650142), vol. 165, pp. 68–76.

59. N. C. for B. I. Bethesda (MD): National Library of Medicine (US), National Center for Biotechnology InformationBethesda (MD): National Library of Medicine (US), blastp [Internet], (available at https://blast.ncbi.nlm.nih.gov/Blast.cgi?PAGE=Proteins).

60. F. Maxwell, I. H. Maxwell, L. M. Glode, Cloning, sequence determination, and expression in transfected cells of the coding sequence for the tox 176 attenuated diphtheria toxin A chain. Mol. Cell. Biol. 7, 1576–1579 (1987).

61. M. V Rodnin, M. M. Kashipathy, A. Kyrychenko, K. P. Battaile, S. Lovell, A. S. Ladokhin, Structure of the Diphtheria Toxin at Acidic pH: Implications for the Conformational Switching of the Translocation Domain. Toxins (Basel). 12 (2020), doi:10.3390/toxins12110704.

62. Z. Turgeon, D. White, R. Jørgensen, D. Visschedyk, R. J. Fieldhouse, D. Mangroo, A. R. Merrill, Yeast as a tool for characterizing mono-ADP-ribosyltransferase toxins. FEMS Microbiol. Lett. 300, 97–106 (2009).

63. A. Vaillant, HBsAg, Subviral Particles, and Their Clearance in Establishing a Functional Cure of Chronic Hepatitis B Virus Infection. ACS Infect. Dis. 7, 1351–1368 (2021).

