## Supplementary text-figure-tables for "Targeting hepatitis B vaccine escape using immunogenetics in Bangladeshi infants"

For the UK Biobank, ethical approval was obtained from the North West Multi-centre Research Ethics Committee (<https://www.hra.nhs.uk/about-us/committees-and-services/res-and-recs/search-research-ethics-committees/north-west-haydock/>).

Supplementary Text

***DT haplotype association studies***

Given the complex LD between HLA alleles at this locus, we also used HLA dimers-based tests to better dissect the HLA-DQ alleles signal (**data S5**). That is, we directly regressed the HLA-DQ combinations found in each participant (e.g. *DQA1*02:01* paired with *DQB1*06:01*) on the DT phenotype (see **Methods**). With this approach, we found that the *DQA1*02:01-DQB1*06:01* haplotype (beta: 0.17, se: 0.08, p: 0.04, 95% CI: 0.01 to 0.33) was still associated with higher DT antibodies, but *DQA1*01:03-DQB1*02:02* (beta: 0.20, se: 0.10, p: 0.04, 95% CI: 0.01 to 0.40) was the haplotype with the largest effect size. Interestingly, in the single allele analyses *DQA1*01:03* and *DQB1*02:02* showed opposite effect direction (for *DQA1*01:03*: beta: -0.26, 95% CI: -0.35 to -0.17; and for *DQB1*02:02*: beta: 0.28, 95% CI: 0.18 to 0.37), which again suggests that LD was biasing our single allele association results.

***The HLA and lack of diphtheria VEMs***

Finally, we performed a similar analysis as above for DT, where we compared the best and the worst HLA-DR and HLA-DQ dimers (**fig. S11**). There were two regions of DT’s toxin A that showed a better binding in the worst dimers and could therefore be a locus of VEMs (similar to the HBV results above): amino-acids 119 to 137, and amino acids 142 to 161. From the used Uniprot sequences, we found only three polymorphisms (T120S, E121K, and E158A), none of which were ever thoroughly studied or described as VEMs in the literature. We therefore used protein BLAST(*59*) on the 15-mers proteins in those segments of toxin A to find any evidence of VEMs. We found only two sites for which pathogenicity were studied: G128D(*60*) and E148K/A(*61*, *62*). Of note, amino acid 148 is a known toxin A active site. For both sites, mutations led to decreased DT pathogenicity. This suggests that DT mutations that would be the most prone to VEMs may be associated with decreased pathogenicity, in accordance with the lack of well-described diphtheria VEMs. We propose that these results provide a suitable “negative control” for our HBV results above.

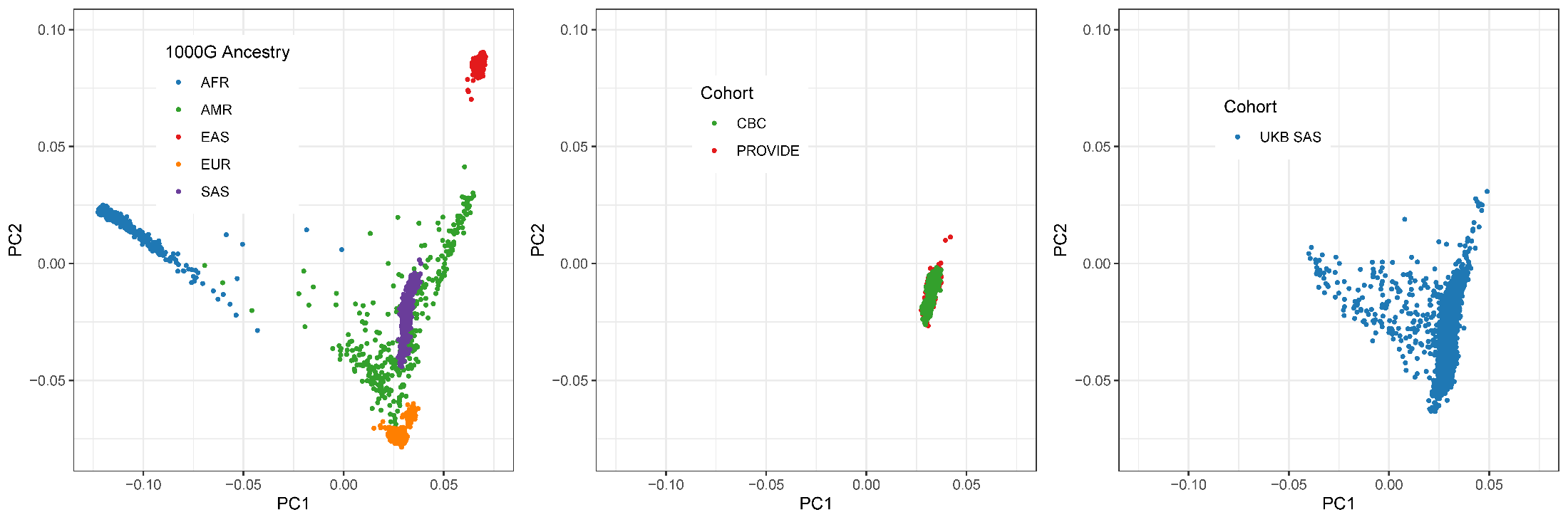

Fig. S1.

Comparison of genetic diversity between the 1000Genome cohort continental genetic ancestries (left), our two Bangladeshi cohorts (middle), and the UKB south Asian genetic ancestry participants who were used as our HLA imputation reference cohort (right). The CBC and PROVIDE cohorts are genetically similar and overlap well with the SAS cohorts in 1000Genome (purple) and in the UKB. AFR: African. AMR: admixed American. EAS: east Asian. EUR: European. UKB SAS: south Asian participants in the UK Biobank. PC1 and PC2: first two genetic principal components (projected on 1000Genome).

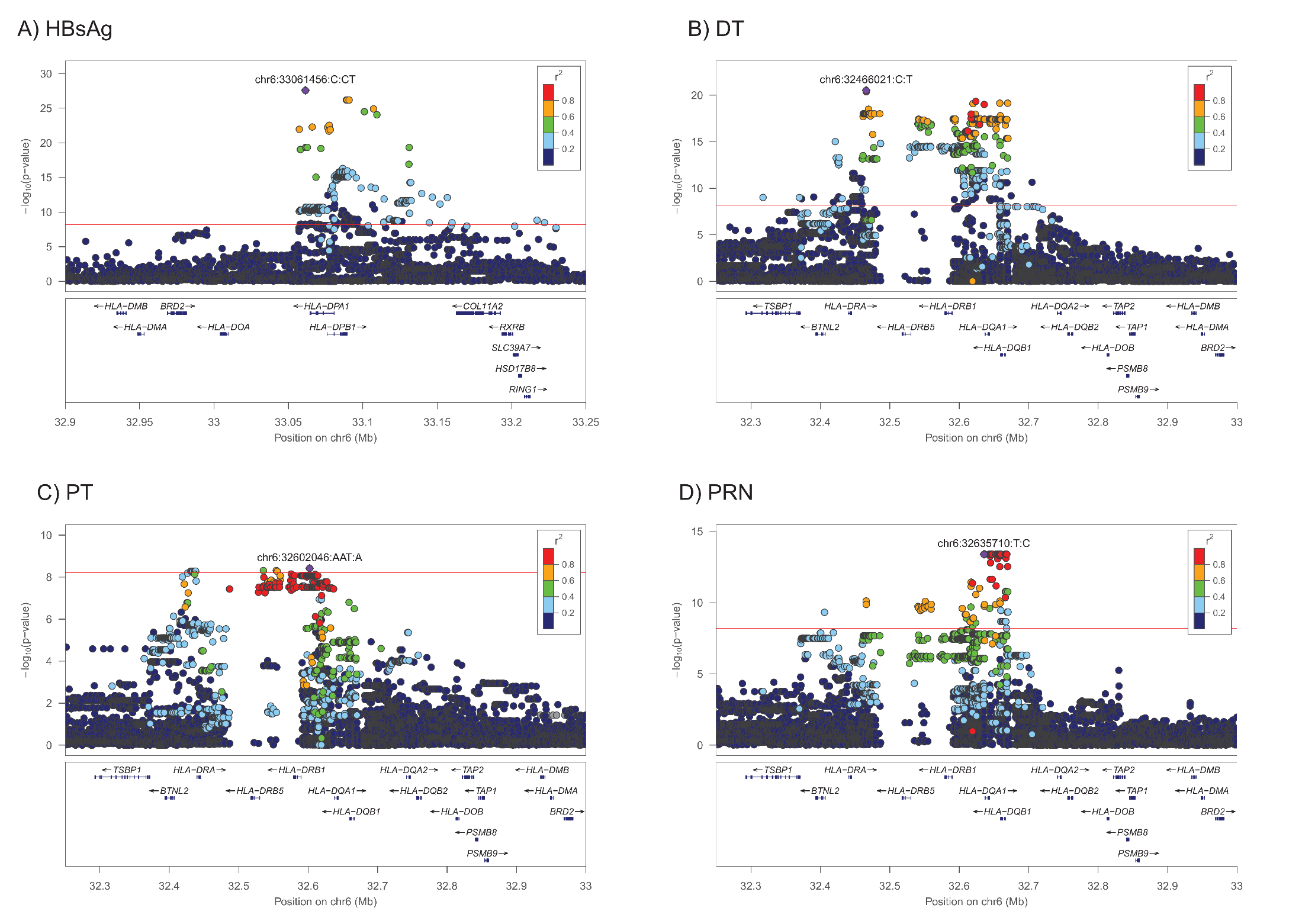

Fig. S2.

Locus zoom plots for the four loci found in **fig. 2**. The purple diamonds with variant annotation show the lead variants from each GWAS. Only protein-coding genes are shown in the bottom panels. Note that since gene tracks are based on the primary GRCh38 contig, *HLA-DRB3* and *HLA-DRB4* are not shown here. They would be in close proximity to *HLA-DRB1* and *HLA-DRB5*. LD was calculated within the cohort directly.

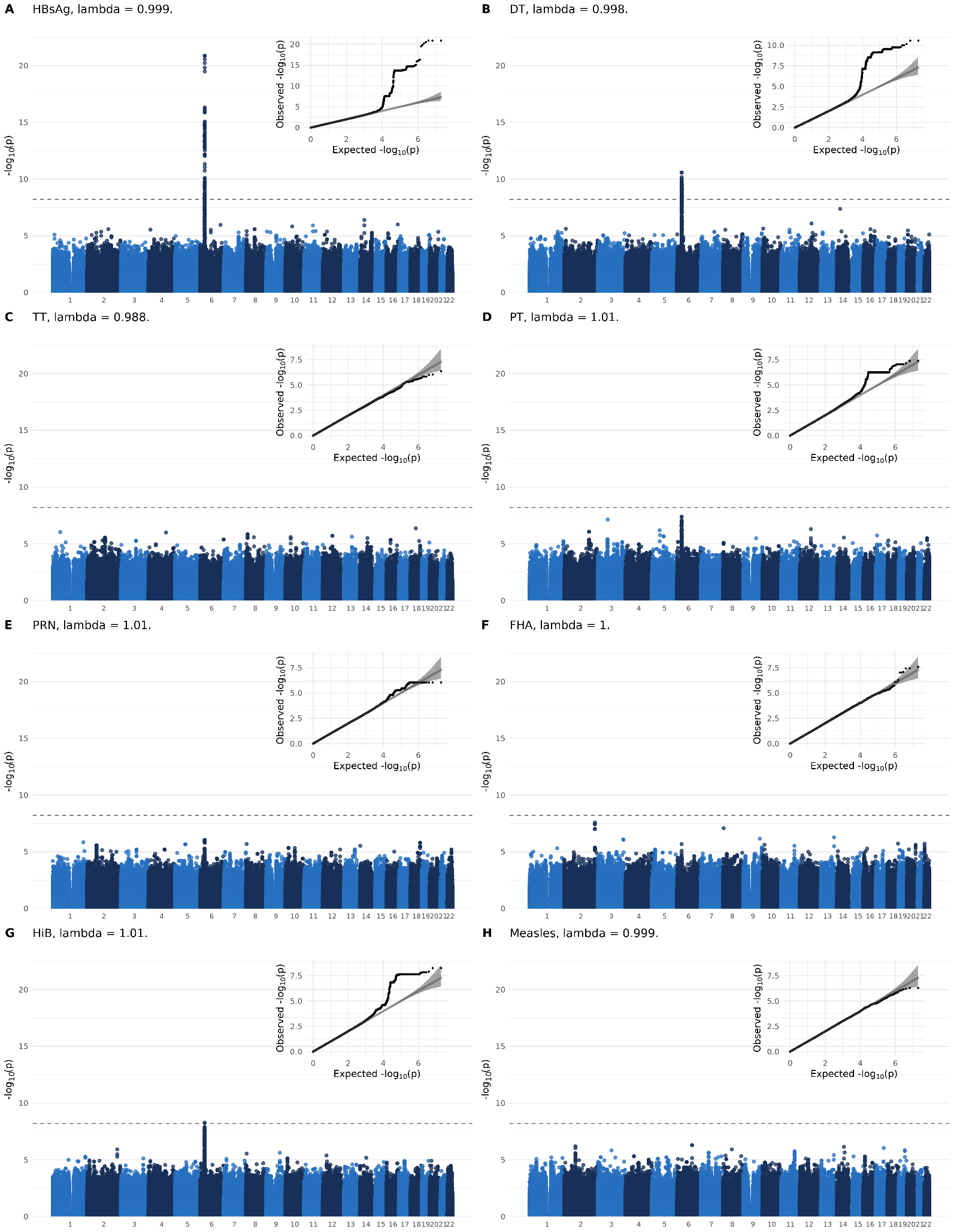

Fig. S3.

Manhattan and qq-plots of the GWAS for each serology phenotype for the CBC cohort only. The genome-wide significance line is set at 5x10^-8^/8.

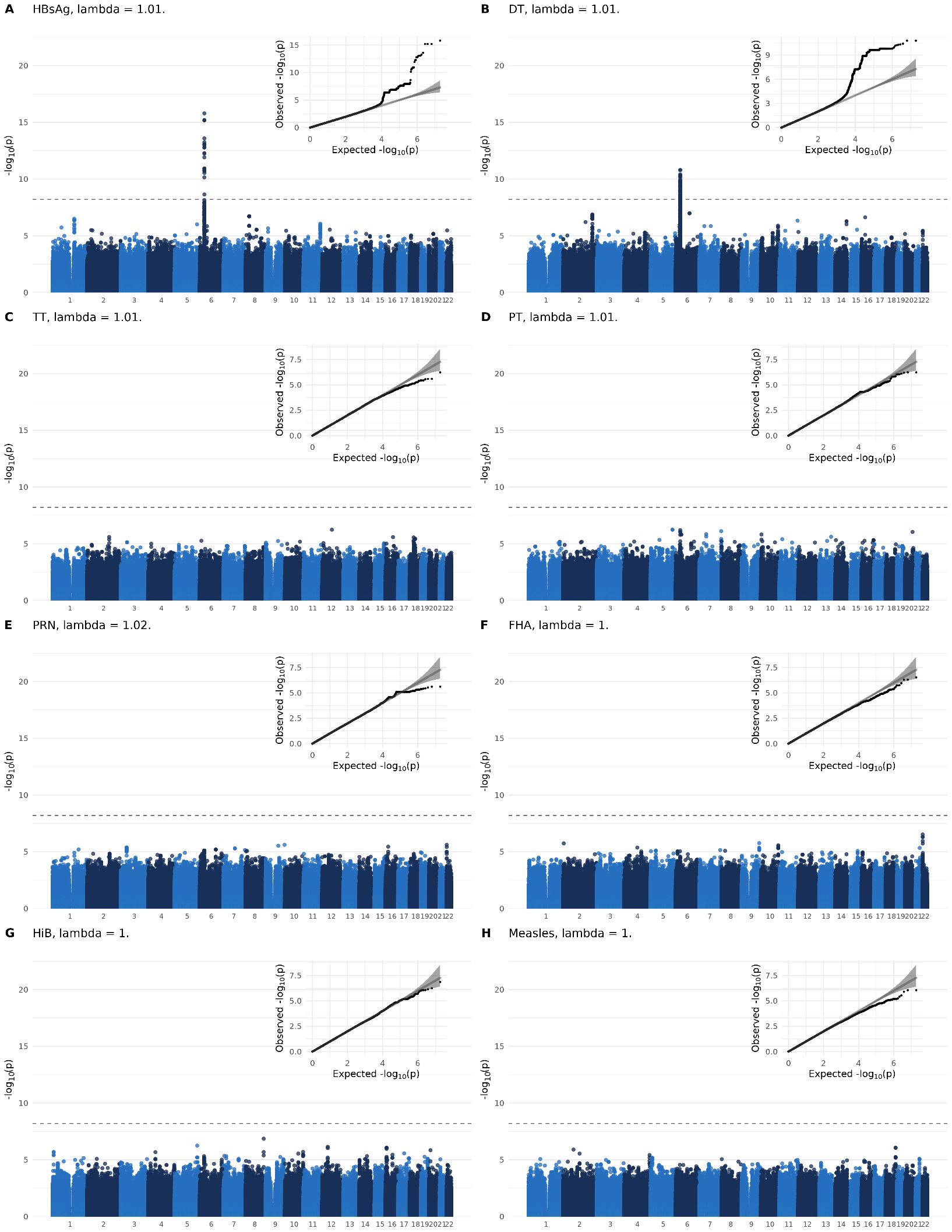

Fig. S4.

Manhattan and qq-plots of the GWAS for each serology phenotype for the PROVIDE cohort only. The genome-wide significance line is set at 5x10^-8^/8.

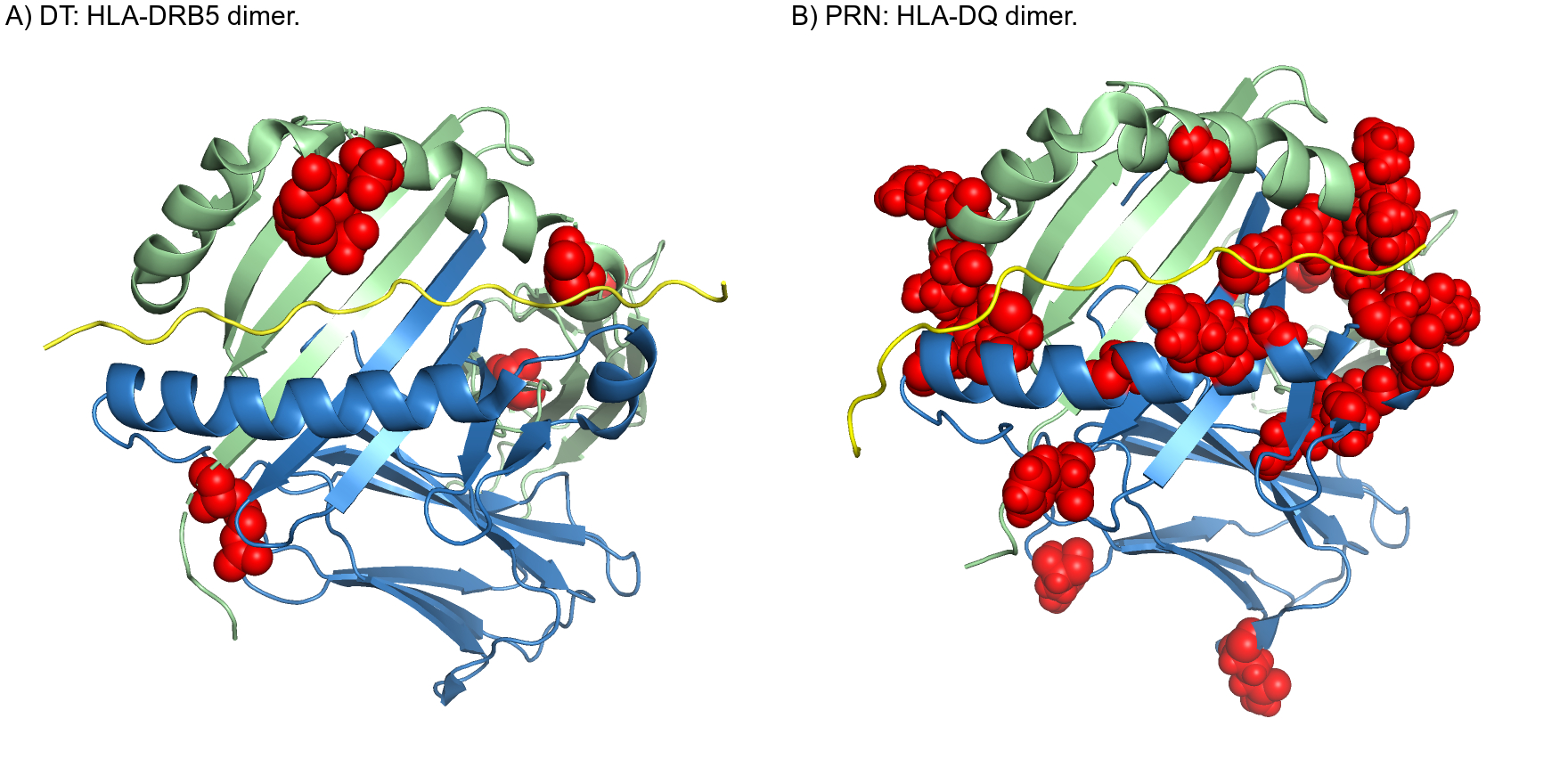

Fig. S5.

Visualization of amino acid residues (in red) associated with each serology phenotype. For each dimer, the blue chains represent the alpha subunits (i.e. DRA, and DQA1), the green chains represent the beta subunits (i.e. DRB1, and DQB1), and the yellow chains show the location of a given epitope in the dimers’ peptide binding grooves.

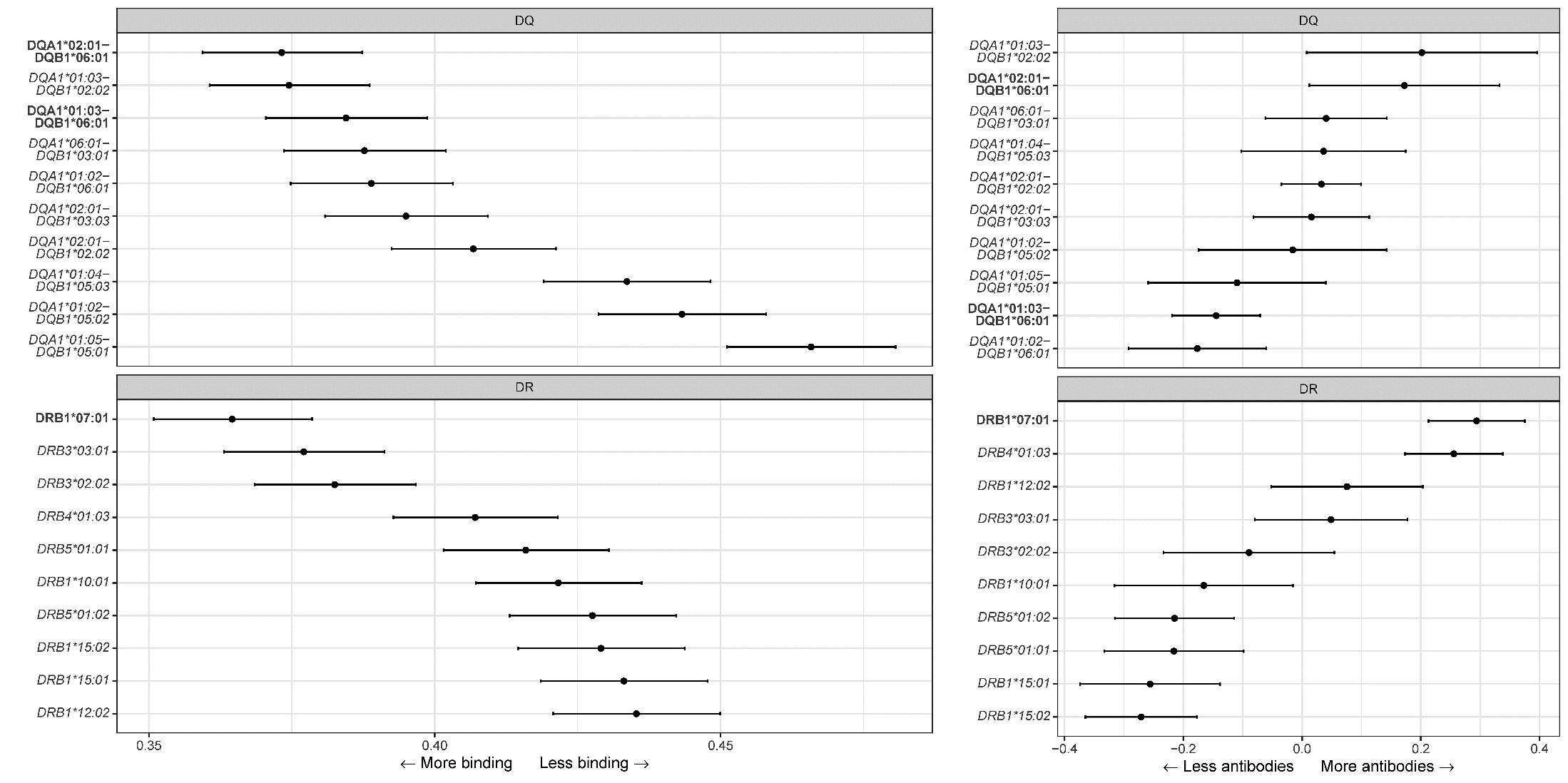

Fig. S6.

*in-silico* binding strength from beta regression (left plots), and results from HLA allele or haplotype association studies (right plots), for the *HLA-DQ* (top) and the *HLA-DR* (bottom) protein dimers for DT. Results are shown as effect estimates and their 95% confidence intervals. Note that for the HLA-DR dimer, the alpha chain comes from the *HLA-DRA* gene which shows almost no polymorphism in humans. The alleles in bold correspond to the alleles with the highest antibodies in the HLA allele association studies. Spearman correlation between estimated antibody levels and relative epitope binding strength: rho = -0.50, p = 0.032. See main text for more details.

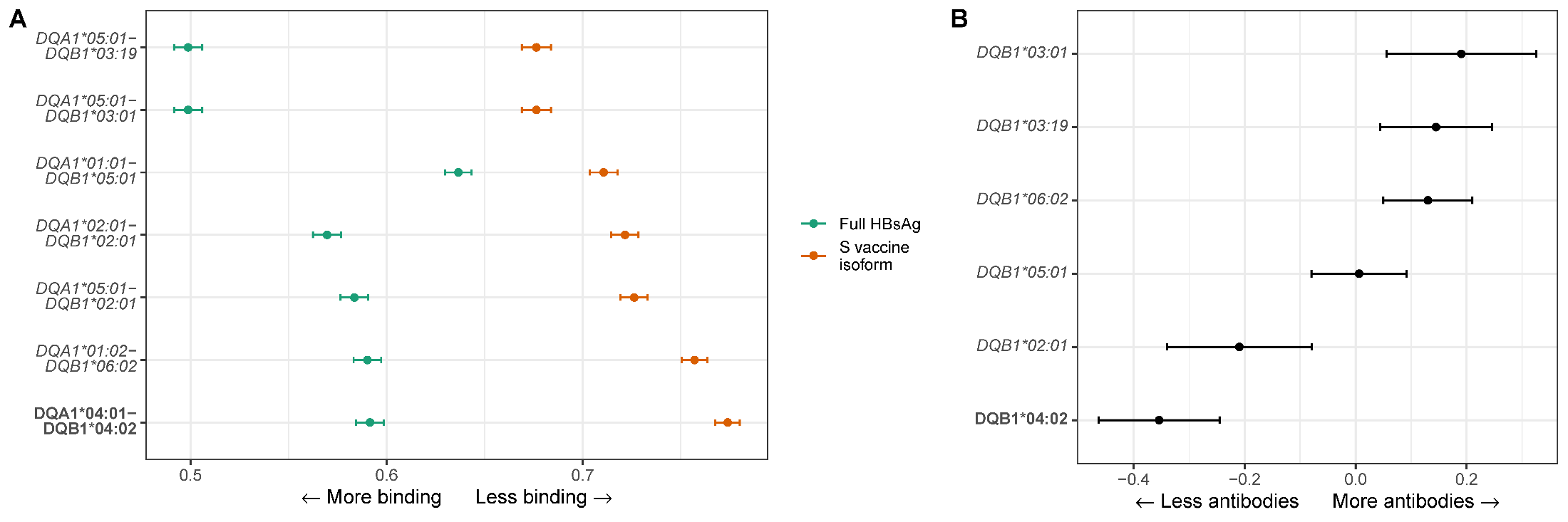

Fig. S7.

*in-silico* binding strength from beta regression (left plot), and results from HLA allele or association studies from the VaccGene sub-Saharan African cohorts (right plot) for HBsAg. Results are shown as effect estimates and their 95% confidence intervals. *HLA-DQA1-DQB1* alleles were chosen to be in LD (r^2^ > 0.2). The alleles in bold correspond to the most significant associations found in the VaccGene study.

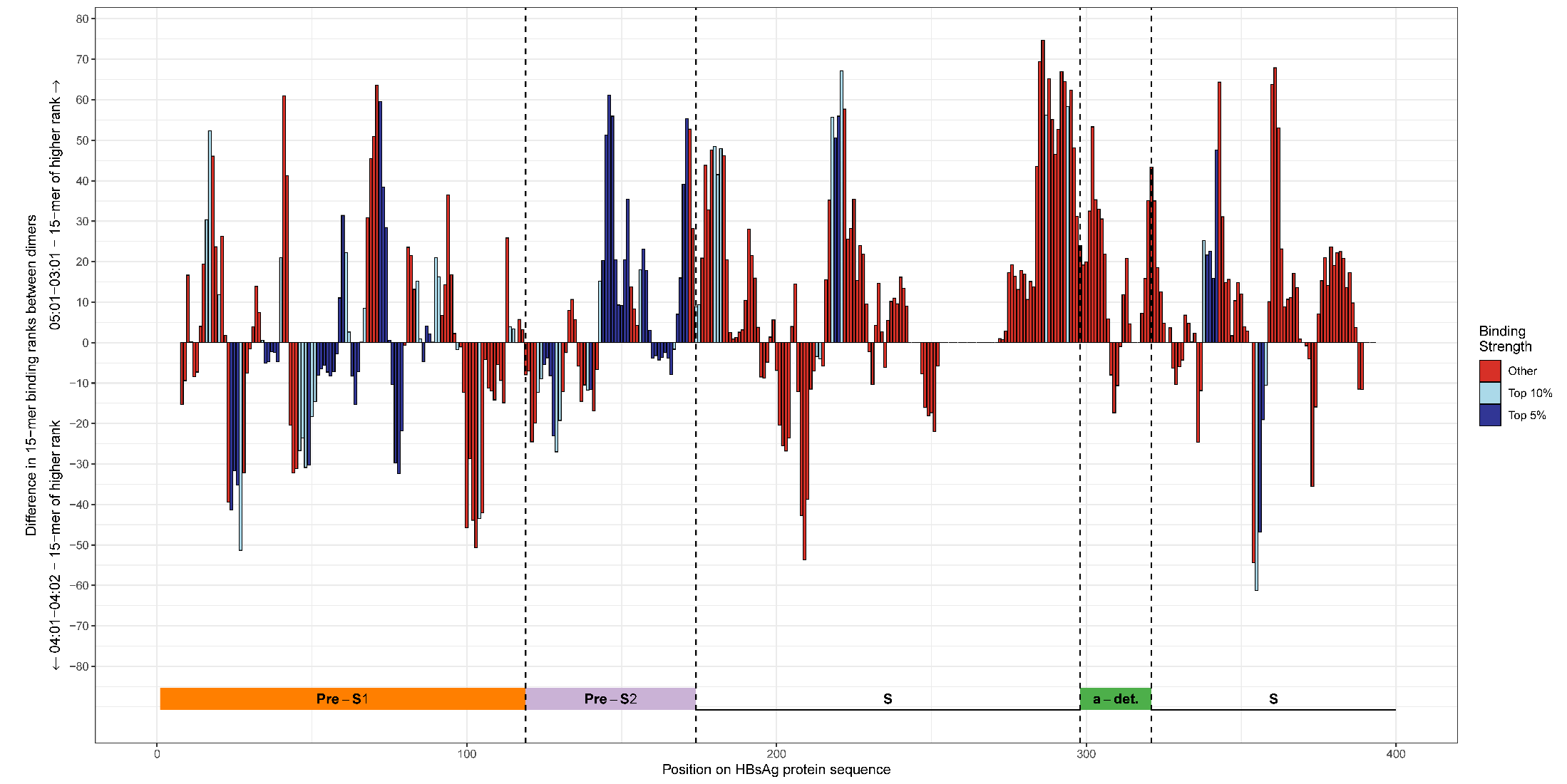

Fig. S8.

Difference in HBsAg binding affinity between the dimer with highest predicted binding affinity to HBsAg peptides (DQA1*05:01-DQB1*03:01) and the dimer with the lowest (DQA1*04:01-DQB1*04:02). The x-axis represents bins of 1 amino-acid across the entire HBsAg sequence. The y-axis represents the difference in binding affinity between the top-binding 15-mers binding in this region for both dimers. Positive values indicate greater binding for DQA1*05:01-DQB1*03:01 than for DQA1*04:01-DQB1*04:02. Negative values indicate the opposite. The colours of the vertical bars refer to the rank of affinity of binding of the 15-mer to either DQA1*05:01-DQB1*03:01 (positive values) or DQA1*04:01-DQB1*04:02 (negative values). Small blue bars mean that both dimers are predicted to bind with high affinity (top 10% quantile) at this position. Long red bars mean that the difference in dimer binding to 15-mer is predicted to be large, but weak for both. A long dark blue bar means that the expected binding difference is large and that one of the dimers bind strongly to the 15-mer. Bottom colored horizontal bar indicates the different section of the HBsAg protein.

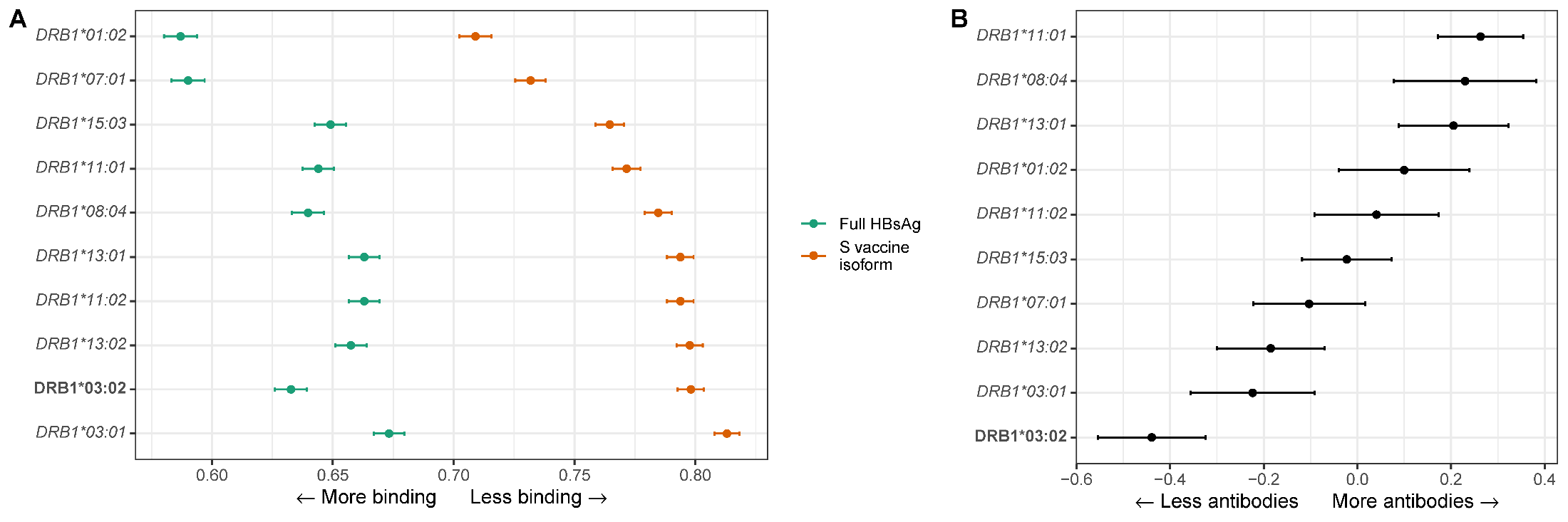

Fig. S9.

*in-silico* binding strength from beta regression (left plot), and results from HLA allele or haplotype association studies from the VaccGene sub-Saharan African cohorts (right plot) for HBsAg. Results are shown as effect estimates and their 95% confidence intervals. Note that for the HLA-DR dimer, the alpha chain comes from the *HLA-DRA* gene which shows almost no polymorphism in humans. The alleles in bold correspond to the most significant associations found in the VaccGene study.

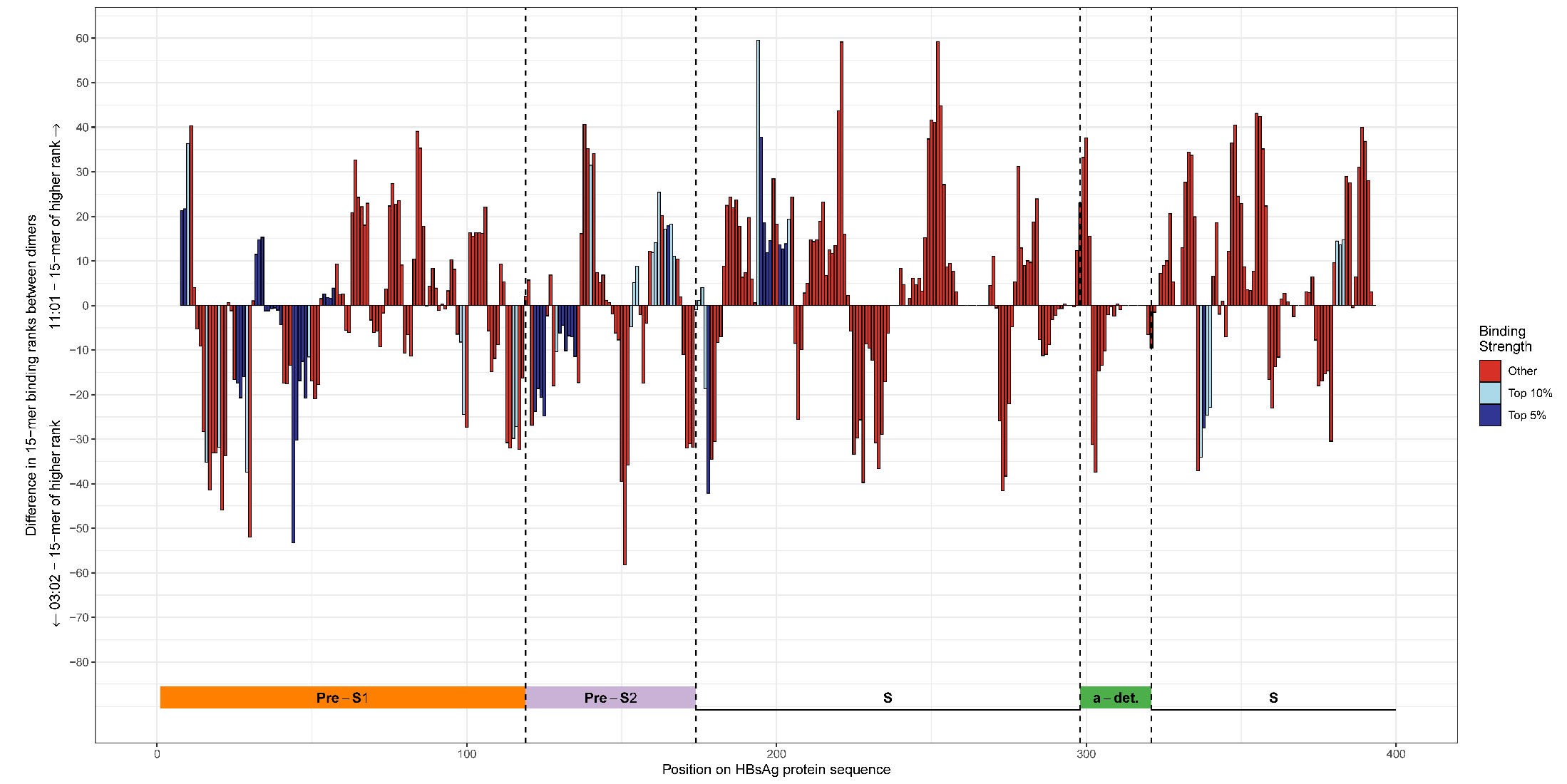
Fig. S10.

Difference in HBsAg binding affinity between the dimer with highest predicted binding affinity to HBsAg peptides (DQA1*05:01-DQB1*03:01) and the dimer with the lowest (DQA1*04:01-DQB1*04:02). The x-axis represents bins of 1 amino-acid across the entire HBsAg sequence. The y-axis represents the difference in binding affinity between the top-binding 15-mers binding in this region for both dimers. Positive values indicate greater binding for DRA*01:01-DRB1*11:01 than for DRA*01:01-DRB1*03:02. Negative values indicate the opposite. The colours of the vertical bars refer to the rank of affinity of binding of the 15-mer to either DRA*01:01-DRB1*11:01 (positive values) or DRA*01:01-DRB1*03:02(negative values). Small blue bars mean that both dimers are predicted to bind with high affinity (top 10% quantile) at this position. Long red bars mean that the difference in dimer binding to 15-mer is predicted to be large, but weak for both. A long dark blue bar means that the expected binding difference is large and that one of the dimers bind strongly to the 15-mer. Bottom colored horizontal bar indicates the different section of the HBsAg protein.

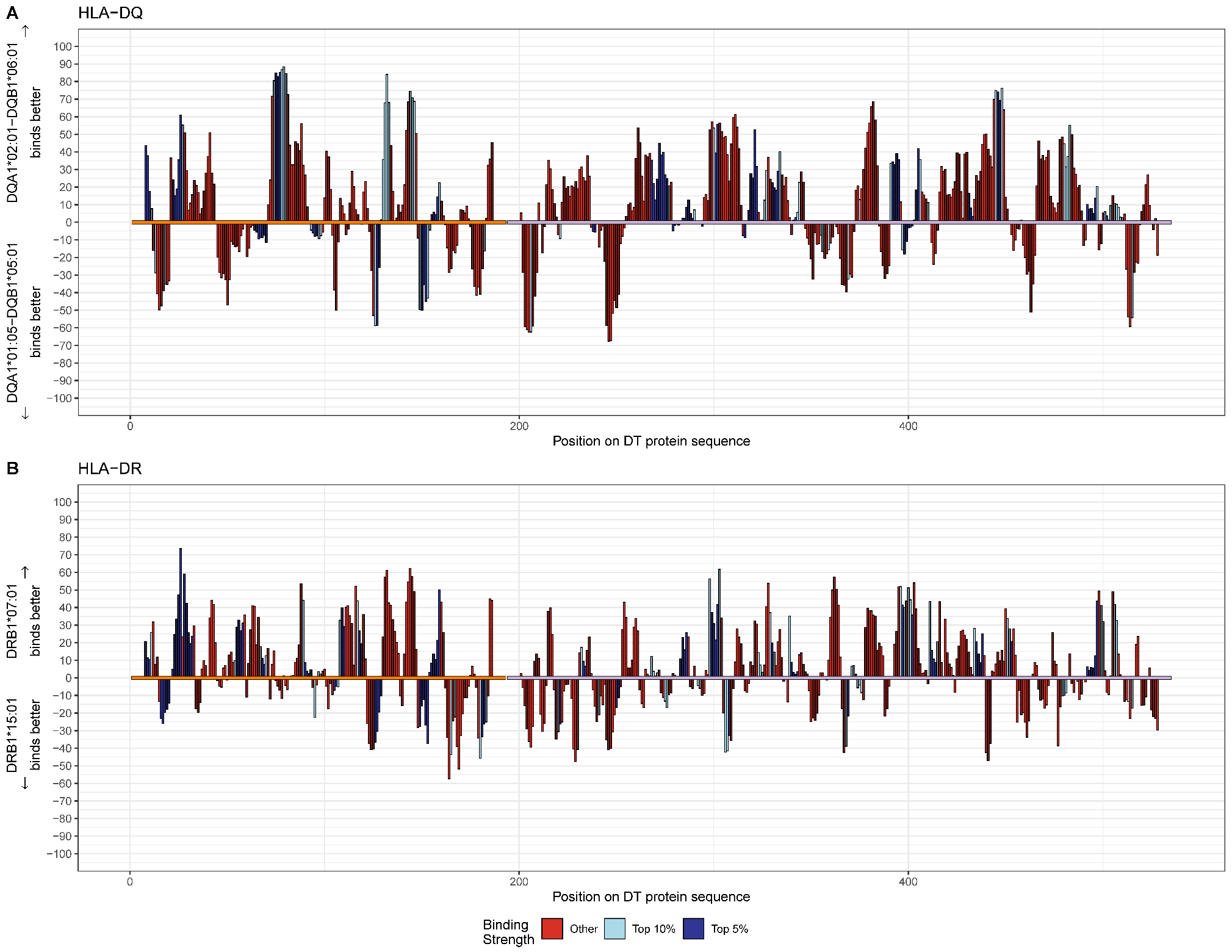

Fig. S11.

Difference in binding strength quantiles between the best binding dimers (DQA1*02:01-DQB1*06:01 and DRB1*07:01) and the worst binding dimer (DQA1*01:05-DQB1*05:01 and DRB1*15:01) for DT. The more positive the difference is, the more epitopes at this region of DT are expected to bind to the best dimer. Negative values mean the opposite. The colours of the bars refer to how strong the binding is to either the best dimer (positive values) or the worst dimer (negative values). Hence, small dark blue bars mean that both dimers bind strongly (top 5% quantile) to the epitopes in this area. Long red bars mean that the difference in dimer binding is expected to be large, but weak for both. A long dark blue bar means that the expected binding difference is large and that one of the dimers bind strongly to the epitopes. The orange bar corresponds to the A toxin, and the purple bar to the B toxin. See main text for interpretation of results.

| **Gene** | **Concordance** |
| --- | --- |
| *HLA-A* | 96.2% |
| *HLA-B* | 93.7% |
| *HLA-DQA1* | 90.3% |
| *HLA-DQB1* | 93.9% |
| *HLA-DRB1* | 96.4% |
| *HLA-DRB3* | 89.6% |
| *HLA-DRB4* | 86.9% |
| *HLA-DRB5* | 89.9% |

Table S1.

Concordance at G-group resolution of our south Asian genetic ancestry imputation panel and 454 sequencing HLA genotyping in a subset of 541 participants.

| **Primary analysis** | | | | | | | |
| --- | --- | --- | --- | --- | --- | --- | --- |
| **Trait** | **Allele** | **Allele conditioned on** | **Effect size** | **Standard error** | **P-value** | **95% Confidence Interval** | **Allele frequency** |
| HBsAg | *DPB1*04:01* | - | 0.481 | 0.042 | 4.53x10^-30^ | (0.399, 0.564) | 31.5% |
| DT | *DRB1*07:01* | - | 0.294 | 0.041 | 1.21x10^-12^ | (0.213, 0.375) | 25.3% |
| DT | *DQA1*02:01* | - | 0.291 | 0.041 | 1.89x10^-12^ | (0.21, 0.373) | 25.4% |
| DT | *DQB1*06:01* | - | -0.312 | 0.047 | 2.50x10^-11^ | (-0.403, -0.22) | 18.9% |
| DT | *DRB4*01:03* | - | 0.256 | 0.042 | 1.18x10^-9^ | (0.173, 0.338) | 24.8% |
| **Conditional analysis** | | | | | | | |
| **Trait** | **Allele** | **Allele conditioned on** | **Effect size** | **Standard error** | **P-value** | **95% Confidence Interval** | **Allele frequency** |
| DT | *DRB1*07:01* | *DQB1*06:01* | 0.235 | 0.042 | 2.36x10^-8^ | (0.152, 0.317) | 25.3% |
| DT | *DQA1*02:01* | *DQB1*06:01* | 0.233 | 0.042 | 2.93x10^-8^ | (0.151, 0.315) | 25.4% |
| DT | *DQB1*06:01* | *DQA1*02:01* | -0.245 | 0.047 | 1.74x10^-7^ | (-0.337, -0.153) | 18.9% |
| DT | *DQB1*06:01* | *DRB4*01:03* | -0.249 | 0.048 | 1.82x10^-7^ | (-0.343, -0.155) | 18.9% |
| DT | *DQB1*06:01* | *DRB1*07:01* | -0.244 | 0.047 | 1.83x10^-7^ | (-0.336, -0.153) | 18.9% |
| DT | *DRB1*07:01* | *DRB4*01:03* | 0.230 | 0.049 | 3.21x10^-6^ | (0.133, 0.326) | 25.3% |
| DT | *DQA1*02:01* | *DRB4*01:03* | 0.228 | 0.049 | 4.01x10^-6^ | (0.131, 0.325) | 25.4% |
| DT | *DRB4*01:03* | *DQB1*06:01* | 0.180 | 0.043 | 2.95x10^-5^ | (0.095, 0.264) | 24.8% |
| DT | *DRB4*01:03* | *DQA1*02:01* | 0.135 | 0.050 | 6.57x10^-3^ | (0.038, 0.233) | 24.8% |
| DT | *DRB4*01:03* | *DRB1*07:01* | 0.131 | 0.050 | 8.04x10^-3^ | (0.034, 0.228) | 24.8% |
| DT | *DRB1*07:01* | *DQA1*02:01* | 0.310 | 0.364 | 3.94x10^-1^ | (-0.402, 1.023) | 25.3% |
| DT | *DQA1*02:01* | *DRB1*07:01* | -0.034 | 0.363 | 9.25x10^-1^ | (-0.746, 0.678) | 25.4% |

Table S2.

HLA allele association studies (top) and conditional analyses (bottom) results. Only HLA alleles which were genome-wide significant (p<5x10^-8^/8) in the primary analyses are shown. For the DT trait, given the multiple significant results in the DQ-DRB haplotype, we show results when conditioning on the significant results from the primary analyses. Rows are ordered by descending p-values. See **fig. 3** for single variant Manhattan plots.

| **Allele** | **CBC+PROVIDE (Bangladesh)** | | | | **VaccGene (Burkina Faso + South Africa + Uganda)** | | | |
| --- | --- | --- | --- | --- | --- | --- | --- | --- |
|  | **Effect (95% CI)** | **Standard error** | **P-value** | **Allele frequency** | **Effect (95% CI)** | **Standard error** | **P-value** | **Allele frequency** |
| *DPB1*04:01* | 0.48 (0.40 to 0.56) | 0.04 | 4.5x10^-30^ | 31.5% | 0.27 (0.12 to 0.50) | 0.08 | 6.3x10^-4^ | 4.3% |
| *DPB1*01:01* | -0.44 (-0.69 to -0.18) | 0.13 | 6.9x10^-4^ | 2.2% | -0.23 (-0.30 to -0.16) | 0.04 | 1.1x10^-10^ | 30.0% |
| *DRB1*03:02* | Not found in cohort | | | | -0.44 (-0.55 to -0.32) | 0.06 | 7.0x10^-14^ | 8.2% |
| *DQB1*04:02^†^* | -0.06 (-0.45 to 0.32) | 0.20 | 0.74 | 1.0% | -0.35 (-0.46 to -0.25) | 0.06 | 1.9x10^-10^ | 9.4% |
| *DQA1*04:01^†^* | Not found in cohort | | | | -0.30 (-0.40 to -0.20) | 0.05 | 5.7x10^-9^ | 11.3% |

Table S3.

Comparison of 2-field HLA allele associations results for the HBsAg phenotype between CBC+PROVIDE and the VaccGene cohort. Differences in allele frequencies are largely responsible for the discrepancy between the results, but our *HLA-DPB1* findings are replicated in VaccGene. ^†^DQA1*04:01 and DQB1*04:02 were in linkage disequilibrium in the VaccGene cohort (r^2^ = 66%).
